# Characterizing and Predicting Post-Acute Sequelae of SARS CoV-2 infection (PASC) in a Large Academic Medical Center in the US

**DOI:** 10.1101/2022.10.21.22281356

**Authors:** Lars G. Fritsche, Weijia Jin, Andrew J. Admon, Bhramar Mukherjee

## Abstract

**Objective:** A growing number of Coronavirus Disease-2019 (COVID-19) survivors are affected by Post-Acute Sequelae of SARS CoV-2 infection (PACS). Using electronic health records data, we aimed to characterize PASC-associated diagnoses and to develop risk prediction models.

**Methods:** In our cohort of 63,675 COVID-19 positive patients, 1,724 (2.7 %) had a recorded PASC diagnosis. We used a case control study design and phenome-wide scans to characterize PASC-associated phenotypes of the pre-, acute-, and post-COVID-19 periods. We also integrated PASC-associated phenotypes into Phenotype Risk Scores (PheRSs) and evaluated their predictive performance.

**Results:** In the post-COVID-19 period, known PASC symptoms (e.g., shortness of breath, malaise/fatigue) and musculoskeletal, infectious, and digestive disorders were enriched among PASC cases. We found seven phenotypes in the pre-COVID-19 period (e.g., irritable bowel syndrome, concussion, nausea/vomiting) and 69 phenotypes in the acute-COVID-19 period (predominantly respiratory, circulatory, neurological) associated with PASC. The derived pre- and acute-COVID-19 PheRSs stratified risk well, e.g., the combined PheRSs identified a quarter of the COVID-19 positive cohort with an at least 2.9-fold increased risk for PASC.

**Conclusions:** The uncovered PASC-associated diagnoses across categories highlighted a complex arrangement of presenting and likely predisposing features, some with a potential for risk stratification approaches.

**Graphical Abstract:** 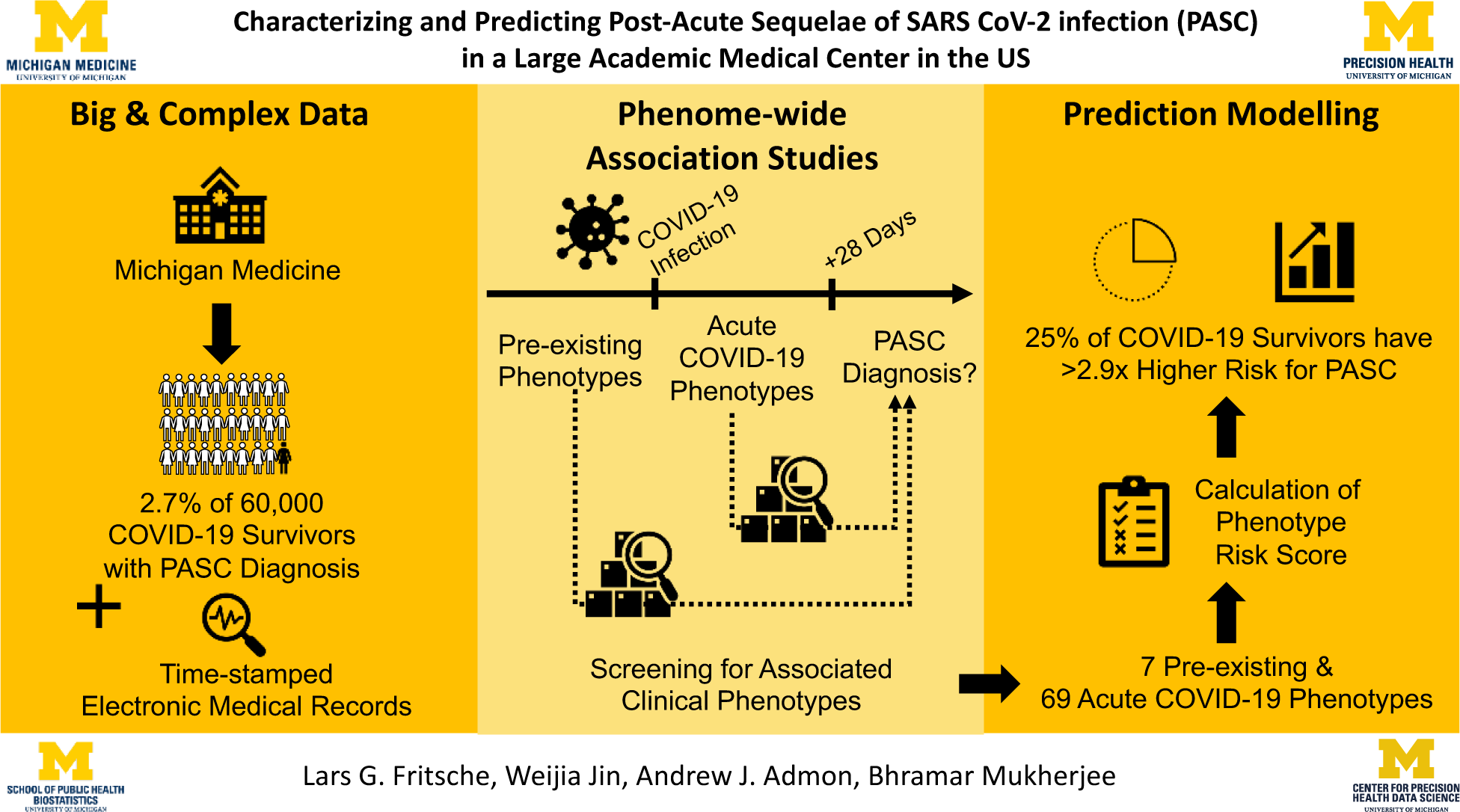

## 1. Introduction

Coronavirus Disease-2019 (COVID-19) has posed unprecedented challenges to the public health and healthcare system. As of September 30, 2022, 96,158,524 confirmed COVID-19 cases were in the US [1]. Studies suggest that 20 to 40% of COVID-19 survivors may be affected by Post-Acute Sequelae of COVID-19 (PASC) [2-4] — also termed Post COVID conditions (PCC), [5, 6], Long COVID [7], Post-Acute COVID-19 Syndrome (PACS) [8], Chronic COVID-19 Syndrome [9], and Long Haul COVID-19 [10]. PASC is an aggregate term for a highly heterogeneous group of post-COVID-19 problems, including persistent symptoms of acute infection (e.g., cough, fatigue, loss of smell [11-13]), new chronic disorders, (e.g., chronic lung or neurologic disease [3, 14-21]), and late post-COVID complications (e.g., autoimmune complications). COVID-19 vaccinations could decrease the risk for PASC by 13% - 22% [22, 23]; however, with a massive number of breakthrough infections and a relaxation of mitigation measures throughout the world, the high prevalence of PASC during an ongoing pandemic could present a tremendous burden for healthcare systems worldwide.

Several demographic factors, preexisting conditions, and biomarkers have been associated with PASC. For example, severe acute COVID-19, female gender, older age, pre-existing diabetes, or the experience of specific symptoms during the acute COVID-19 phase, including fatigue, headache, hoarse voice, etc., were reported to increase the risk for PASC [24-27]. Carlo et al. reported an immunoglobulin (Ig) signature, based on total IgM and IgG3, as a predictor for PASC [28]. Emily et al. identified a series of features, including the rate of healthcare utilization, patient age, dyspnea, and other diagnosis and medication information, to predict PASC [29]. In Su et al.’s study, four risk factors: type 2 diabetes, SARS-CoV-2 RNAemia, Epstein-Barr virus viremia, and specific auto-antibodies were identified [30].

Together, these studies highlight the possibility and the need to uncover and understand PASC risk factors to identify and protect vulnerable groups. Furthermore, a better understanding of PASC might allow the identification of PASC subtypes and their specific risk profiles. However, the novelty of this condition and the sparsity of studies so far have hampered the development of risk-prediction models for PASC.

In our current study, we aim to fill this gap by identifying predisposing diagnoses of PASC through phenome-wide association studies (PheWAS) of the pre-COVID-19 and acute-COVID-19 time periods and then use the identified pre-existing conditions to develop and evaluate integrated and usable Phenotype Risk Scores (PheRS) [31] to predict PASC [32, 33]. To do this, we leverage a cohort of over 60,000 COVID-19-positive patients cared for at Michigan Medicine (MM), a large academic medical center in the Midwestern US, between March 2020 and August 2022. This cohort includes 1,724 patients that were subsequently diagnosed with PASC using diagnostic codes or clinical problem lists. With its rich retrospective EHR data that includes socioeconomic status (SES), demographics, and other relevant variables, this cohort offers a unique opportunity to study PASC.

## 2. Subjects and Methods

### 2.1. Study cohort

The study included Michigan Medicine (MM) patients with a recorded COVID-19 diagnosis or a positive real-time reverse transcriptase chain (RT-PCR) test for SARS-CoV-2 infection performed/recorded at MM between March 10, 2020, and August 31, 2022. Diagnoses were recorded at clinic visits and hospital encounters. RT-PCR testing data was collected for routine screening at hospital admission, before procedures, and for employee screening. Tests included both symptomatic and asymptomatic individuals.

For each subject, the date of their first COVID-19 diagnosis or RT-PCR positive test, whichever came first, was considered the index date. Dates were regarded as protected health information and operationalized as days since birth; however, the quarter of the year of the index date was obtained. To allow sufficient follow-up time for diagnosing PASC, we limited the analysis to patients with encounters at least two months after being COVID-19 positive and stratified them in PASC cases (had a recorded PASC diagnosis) and PASC controls (had no recorded PASC diagnosis).

PASC diagnoses were either based on an entry of PASC in the diagnosis section of the EHR database’s Problem Summary List (PSL, **Table S1**) or on observations of the ICD-10-CM (International Classification of Diseases codes, tenth edition with clinical modifications) U09.9 (“Post COVID-19 condition, unspecified”) or B94.8 (“Sequelae of other specified infectious and parasitic diseases”). The CDC recommended the latter as a temporary alternative to the PASC-specific U09.9 code, which was implemented on October 1, 2021 [34]. PSL diagnoses represent active and resolved patient problems entered by healthcare providers. The age at the first observed ICD- or PSL-based PASC diagnosis was considered the age of onset of PASC. PASC cases (see definition below) without a prior positive test were excluded because the timepoint of the test was crucial for defining the pre-COVID-19 and acute-COVID-19 time periods (**Figure 1**).

**Figure 1:**
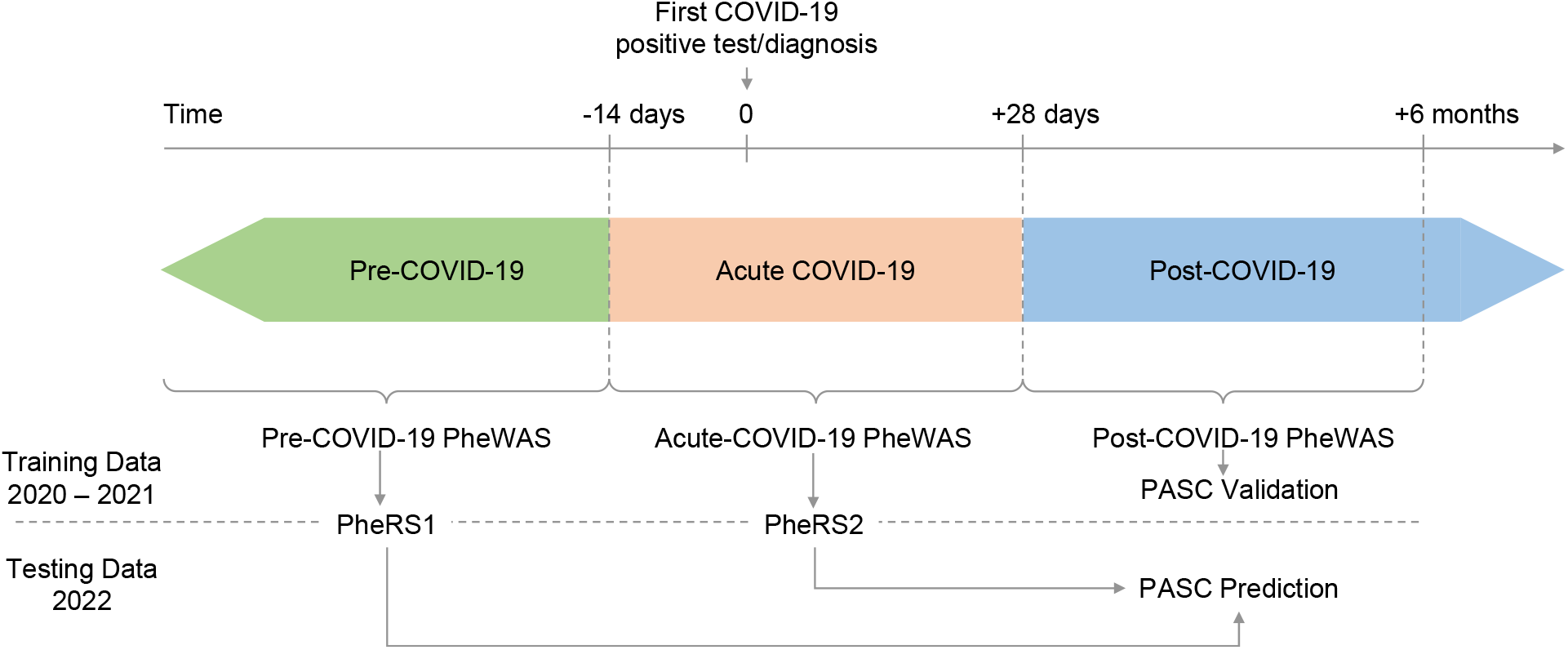
Schematic on study design. Three time periods were defined relative to the 1. positive COVID-19 test or diagnosis (index date): pre-COVID-19 until −14 days, acute-COVID-19 from − 14 to +28 days, and post-COVID-19 from +28 days onwards. The post-COVID-19 PheWAS is used to validate features of PASC cases compared to COVID-19 cases without PASC diagnoses. The Pre-COVID-19 and acute-COVID-19 PheWAS on the training data (index date in 2020 – 2021) inform on phenotype risk scores (PheRS) that will be used to predict PASC in the testing data (index date in 2022).

We also categorized PASC patients based on ICD10 diagnoses concurrently recorded with their first PASC diagnosis and mapped them to 29 phenotype concepts previously reported as common PASC symptoms [3]. In addition, we manually mapped detailed PSL diagnoses to these 29 concepts (**Table S1 and S2**).

### 2.2. Definition of demographics, socioeconomic status, and other covariates

To examine and adjust for confounding by patient characteristics, socioeconomic status, and other variables, we obtained the following data for each participant: age, self-reported gender, self-reported race/ethnicity, Neighborhood Disadvantage Index (NDI) without proportion of Black (coded as quartiles, with larger quartiles representing more disadvantaged communities) [35, 36], and population density measured in persons per square mile (operationalized as quartiles).

Additional covariates included vaccination status, the Elixhauser comorbidity score [37, 38], COVID-19 severity (non-severe [not hospitalized] and severe [hospitalized or deceased]), healthcare worker (HCW) status, the timespan of records in the EHR before and after the COVID-19 test/diagnosis, the timespan of records in the EHR before 2020 (referred to as “pre-pandemic” time period). These timespans were based on the first or last recorded encounter in the EHR data. Additional details and definitions of these covariates can be found in **Text S1** and **Table S3**.

We assumed completely at random missingness of the covariates included in our adjusted analyses and performed complete case analyses for each adjustment.

Ethical review and approval were waived for this study due to its qualification for a federal exemption as secondary research for which consent is not required. Determination for exemption made by the Institutional Review Board of the University of Michigan Medical School (IRBMED; study ID: HUM00180294).

### 2.3. Time-restricted phenomes

We constructed each subject’s medical phenome by extracting available ICD9 and ICD10 codes from the EHR and mapping them to 1,813 broader phenotype concepts (PheCodes) using the R package “PheWAS” [39, 40]. In short, individuals with ICD codes that map to a specific PheCode were coded as “1”, then individuals with ICD codes that map to the PheCode’s specific exclusion criteria were coded as missing, and finally, all remaining individuals were coded as “0” for that particular PheCode (further details are described elsewhere [40]). We created three time-restricted phenomes relative to the index date: post-COVID-19 (+28 days to +6 months), pre-COVID-19 (predating −2 weeks), and acute COVID-19 (−14 and +28 days; **Figure 1**).

### 2.4. Matching

To minimize confounding when we compare PASC (case) versus no PASC (control), we matched each PASC case to up to 10 PASC controls using the R package “MatchIt” [41]. Nearest neighbor matching was applied for age at index date, pre-COVID-19 years in EHR, and post-COVID-19 years in EHR. Exact matching was used for sex, primary care visit at Michigan Medicine within the last two years (yes/no), race/ethnicity, and year quarter of the index date. We retained the case-control matching throughout all analyses.

### 2.5. Statistical analysis

#### 2.5.1. PASC-associated PheCodes in Post COVID-19 Period

To characterize diagnoses enriched in COVID-19 patients with PASC, we also conducted PheWAS to identify phenotypes associated with PASC in the post-COVID-19 period (at least 28 days after the COVID-19 index date, see **Figure 1**) using Firth bias-corrected logistic regression by fitting the following model for each PheCode of the post-COVID-19 period phenome:

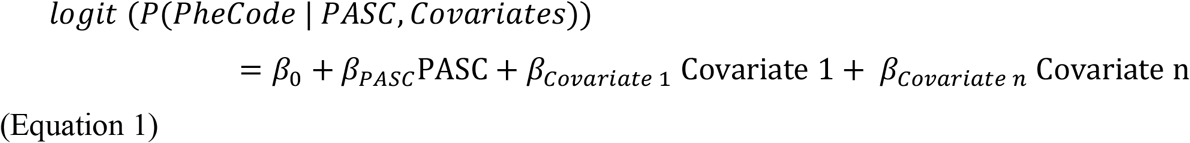

Where covariates were pre-COVID-19 Elixhauser Score (AHRQ), NDI, Population density, healthcare worker status (HCW), vaccination status, and severity, details are summarized in **Text S1 and Table S3**.

#### 2.5.2. Pre-disposing PheCodes

We conducted PheWAS to identify PheCodes pre-disposing to PASC using either PheCodes from the pre-COVID19 period or PheCodes from the acute-COVID-19 period. We ran Firth bias-corrected logistic regression by fitting the following model for each PheCode of the corresponding time-restricted phenome:

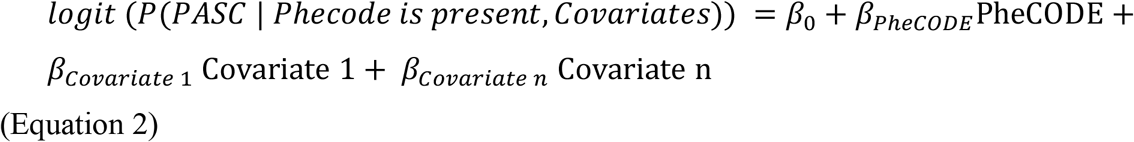

We applied a similar set of covariate adjustments as before (**Table S3**).

The phenomes were split into a training set (index dates in 2020 and 2021) and a testing set (index date in 2022). This choice was to retain the true spirit of future prediction using past data. The training set was used to identify predisposing PheCodes in phenome-wide association studies (PheWAS), while the testing set was used to evaluate prediction models based on the PheWAS results.

To evaluate the robustness of effect sizes of predisposing PheCodes, we performed several sensitivity analyses: (1) females only, (2) males only, (3) index date in 2020, (4) index date in 2021, (5) non-severe outcomes (not hospitalized), (6) severe outcomes (hospitalized or deceased), (7) recorded within two years before the index date, and (8) pre-pandemic (before 2020). For the acute-COVID-19 PheWAS, we excluded PASC cases whose first recorded PASC diagnosis was observed less than 28 days after the index date. The sample sizes of the complete case analyses for various analyses are listed in **Table S4**.

PheWASs were restricted to PheCodes observed at least five times among cases and among controls. For all PheWAS, we excluded PheCode 136 “Other infectious and parasitic diseases” as it included the ICD-10 code “B94.8” which was used to record a PASC diagnosis. For each PheWAS, we applied a Bonferroni correction adjusting for the number of analyzed PheCodes (**Table S4**). In Manhattan plots, we present –log10 (*p*-value) corresponding to tests for association of the underlying phenotype. Directional triangles on the PheWAS plot indicate whether a trait was positively (pointing up) or negatively (pointing down) associated.

We also tested for differences between effect sizes of three subgroup comparisons (non-severe vs. severe outcome, female vs. male, and index date in 2020 vs. 2021) using the following t-statistics:

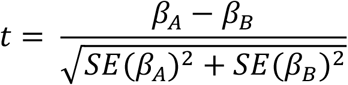

where *β*_*A*_ and *β*_*B*_ are the subgroup-specific beta-estimates with corresponding standard errors *SE*(*β*_*A*_) and *SE*(*β*_*B*_).

#### 2.5.3. Phenotype Risk Scores (PheRS)

##### 2.5.3.1. PheRS Generation

To generate PheRS, we considered two sets of PheCodes: PheCodes that were phenome-wide significant in the pre-COVID-19 PheWAS (considered for the pre-COVID-19 PheRS [PheRS1]) or PheCodes that were phenome-wide significant in the acute-COVID-19 phenome (considered for the acute-COVID-19 PheRS [PheRS2]).

For each of the two sets of PheCodes, we performed ridge penalized logistic regression using the R Package package “glmnet” [42, 43] to obtain the weights per PheCode from the training data before calculating the PheRS as the weighted sum of the presence/absence (coded as 1 and 0) of a PheCode in the testing data.

##### 2.5.3.2. PheRS Evaluation

To evaluate each of the PheRS, we fit the following Firth bias-corrected logistic regression model adjusting for age, gender, race/ethnicity, Elixhauser Score, population density, NDI, HCW, vaccination status, pre-COVID19 years in EHR and severity using a complete case analysis:

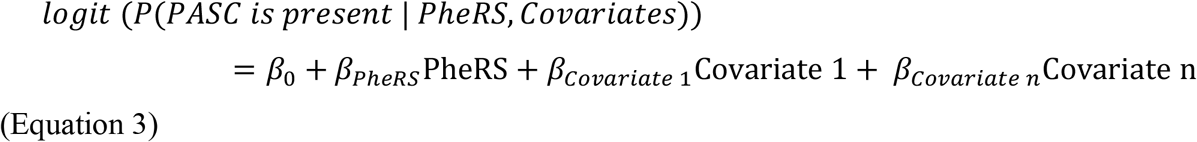

For each PheRS, we assessed the following performance measures relative to the PASC status: (1) overall performance with Nagelkerke’s pseudo-R^2^ using R packages “rcompanion” [44], (2) accuracy with Brier score using R package “DescTools” [45]; and (3) ability to discriminate between PASC cases and matched controls as measured by the area under the covariate-adjusted receiver operating characteristic (AROC; semiparametric frequentist inference) curve (denoted AAUC) using R package “ROCnReg” [46]. Firth’s bias reduction method was used to resolve the problem of separation in logistic regression (R package “brglm2”) [47]

To also evaluate models with both predictors (PheRS1-Ridge + PheRS2-Ridge), we combined them by first fitting a logistic regression with the predictors in the training set to obtain the linear predictors that we used to get the combined score in the testing data.

Unless otherwise stated, analyses were performed using R 4.2.0 [48].

## 3. Results

### 3.1. Patient characteristics

Among 63,675 COVID-19-positive patients who were seen in MM at least two months after their first recorded COVID-19 infection, 1,724 (2.7%) received a PASC diagnosis. The PASC prevalence within three months of a COVID-19 infection ranged from 0.18% (Q3 of 2020) to 1.8% (Q3 of 2021). The most PASC cases were observed in Q4/2021 (n = 134), coinciding with the second peak of positive tests at MM (**Table 1**; **Figure S1**).

**Table 1:**
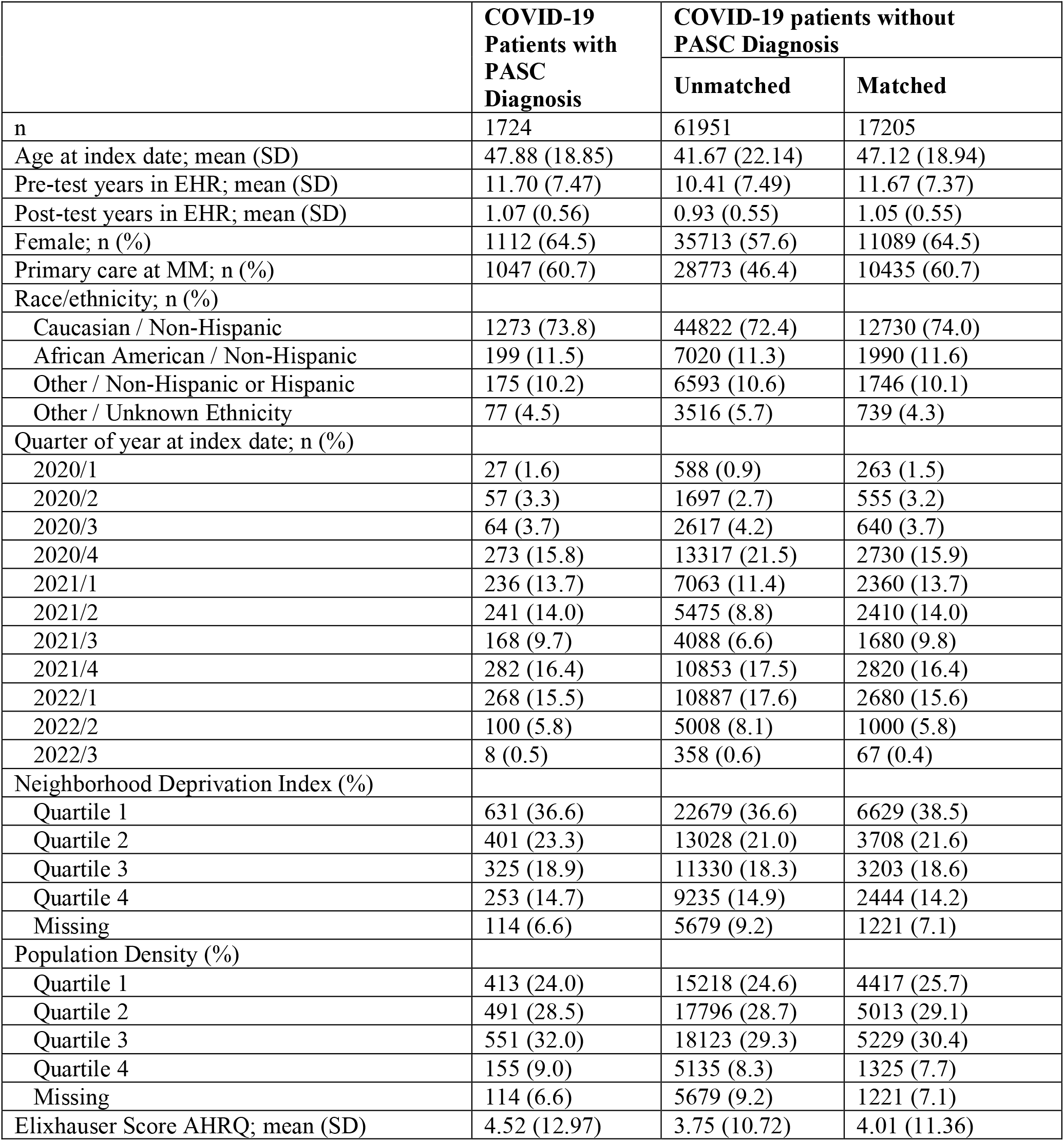
Patient characteristics of COVID-19 patients with (cases) and without observed PASC diagnosis (controls). Case-control matching was based on nearest neighbor matching (age at index date, pre-test years in EHR, post-test years in EHR) and exact matching (gender, primary care at MM, race/ethnicity, quarter of year at COVID-19 index date).

We observed that PASC cases compared to controls were on average older at their index date (mean age 47.9 versus 41.7 years), had a slightly longer timespan covered in the pre-test EHRs (11.7 versus 10.4 years), were more likely female (64.5% versus 56.7%), more likely to have received primary care at MM in the last two years (60.7% versus 46.4%) and showed different distributions across the year quarters over time (**Table 1**).

### 3.2. PASC symptoms / post-COVID-19 PheWAS

When categorizing 1,362 PASC cases with concurrent diagnoses based on 29 previously reported symptoms [3] (362 of the 1,724 cases had no concurrent diagnoses, **Table S1 and S2**), the ten most common diagnoses were: shortness of breath (34.3%), anxiety (30.6%), malaise and fatigue (28.5%), depression (27.2%), sleep disorders (25.4%), asthma (23.6%), headaches (21.4%), migraine (13.8%), cough (13.0%) and joint pain (12.6%) (**Table S5**).

In the post-COVID-19 PheWAS of 1,256 cases versus 12,492 matched controls, all 29 PASC symptoms were enriched among PASC cases (OR > 1), and 27 reached phenome-wide significance (P < 0.05/960 tested PheCodes; P < 5.2e-05) while two were not significant (**Table S6**). In addition to PASC-related phenotypes (e.g., shortness of breath: OR = 9.03 [7.77, 10.50], P = 2.94E-181; malaise and fatigue: OR = 6.17 [5.33, 7.14], P = 2.32E-132; and cardiac dysrhythmias: OR = 2.75 [2.37, 3.18], P = 3.95E-41), many additional diagnoses were enriched in PASC cases, among others musculoskeletal disorders (e.g., costochondritis: OR = 6.88 [95%: 3.05, 14.8], P = 6.72e-08), infectious diseases (e.g., septicemia: OR = 2.31 [1.66, 3.16] P = 2.67e-07), and digestive disorders (e.g., gastroesophageal reflux disease [GERD]: OR = 1.72 [1.50, 1.99], P = 5.10e-14) (**Figure 2, File S1A**).

**Figure 2:**
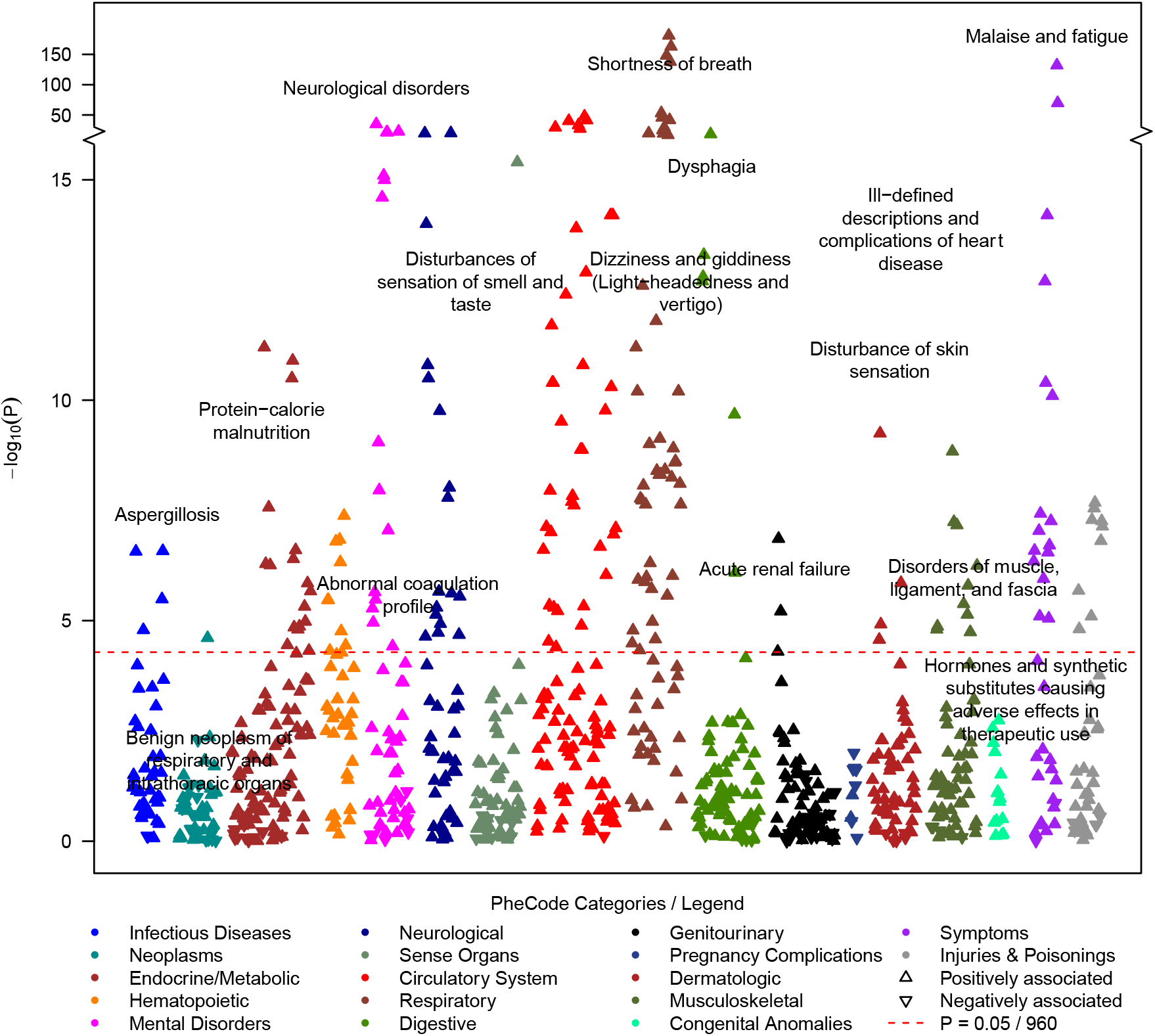
PheWAS on symptoms that occurred between 28 days and 6 months after the first COVID-19 test (Outcome: post-COVID-19 symptoms / phecodes; predictor: PASC diagnosis yes/no). Among PheCodes that reached phenome-wide significance (red dashed line, P <= 0.05/960 = 5.2e-05), only the strongest association per PheCode category was labeled. The analysis was adjusted using the following covariates: age at index date, gender, race/ethnicity, Elixhauser Score AHRQ, population density (quartiles), NDI (quartiles), health care worker status, vaccination status, post-test years in EHR, and severity. Summary statistics can be found in **File S1**.

### 3.3. Pre-COVID-19 PheWAS

To identify potential PASC-predisposing conditions, we performed a PheWAS using the pre-COVID-19 phenome, comparing 1,212 PASC cases versus 11,919 matched controls. Among 1,405 tested PheCodes, seven reached phenome-wide significance (P < 3.56e-05): irritable bowel syndrome (IBS; OR = 1.78 [1.44, 2.18], P = 4.00e-8), concussion (OR = 1.95 [1.51, 2.49], P = 1.24e-07), nausea and vomiting (OR = 1.45 [1.26, 1.67], P = 2.90e-07), shortness of breath (OR = 1.51 [1.29, 1.76] 3.38e-07), respiratory abnormalities (OR = 1.39 [1.22, 1.59], P = 1.10e-06), allergic reaction to food (OR = 1.94 [1.42, 2.60], P = 1.66e-05) and general circulatory disease (OR = 1.52 [1.24, 1.85], P = 3.30e-05; **Figure 3, File S1B**). Additional sensitivity analyses indicated robust associations across various settings (females only, males only, 2020 only, 2021 only, non-severe outcome, severe outcomes, within two years before the index date, or before the pandemic, **Figures S3 A-G, File S1D-F**).

**Figure 3.**
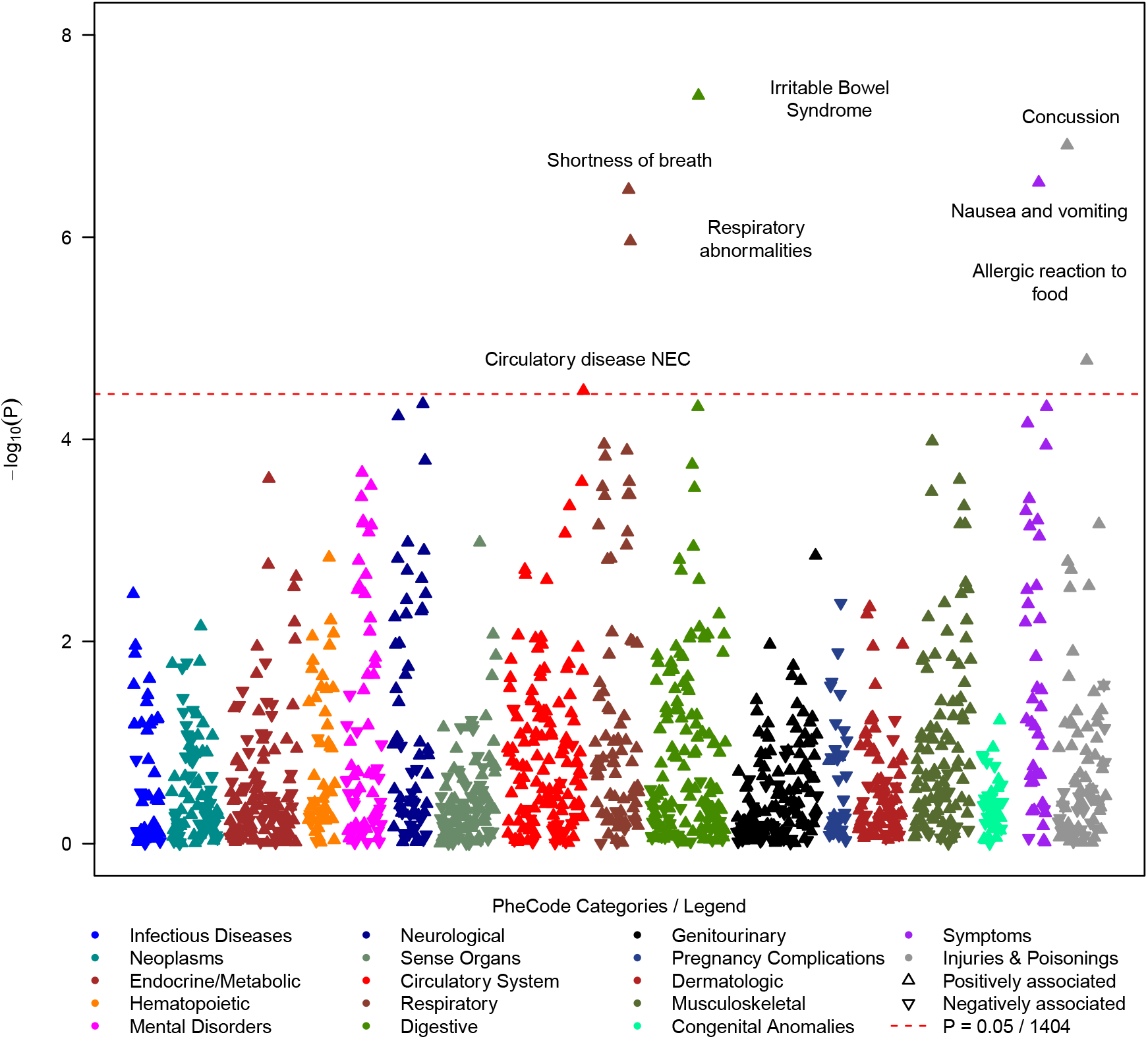
PheWAS on symptoms that occurred at least 14 days before the first positive COVID-19 test (Outcome: PASC diagnosis yes/no; predictors: PheCodes). Among PheCodes that reached phenome-wide significance (red dashed line, P <= 0.05/1404 = 3.56e-05), only the strongest association per PheCode category was labeled. The analysis was adjusted using the following covariates: age at index date, gender, race/ethnicity, Elixhauser Score, population density (quartiles), NDI (quartiles), health care worker status, vaccination status, pre-test years in EHR, and severity. Summary statistics can be found in **File S1**.

### 3.4. Acute-COVID-19 PheWAS

To uncover PASC-predisposing acute-COVID-19 symptoms, we screened 664 phenotypes of the acute-COVID-19 phenome, comparing 874 cases with 8,671 controls. To not identify actual PASC symptoms compared to pre-PASC symptoms, we excluded cases whose PASC diagnosis was recorded less than 28 days after their index date and only retained their matched controls. A total of 69 phenotypes was significantly associated with PASC (P < 7.54e-05) and included, among others, 22 respiratory phenotypes (e.g., shortness of breath, respiratory failure/insufficiency/arrest, dependence on respirator or supplemental oxygen, and cough), 13 circulatory system phenotypes (orthostatic hypotension, hypotension), seven neurological phenotypes (e.g., sleep disorder, migraine, pain), six digestive phenotypes (e.g., GERD, IBS), five mental health phenotypes (e.g., anxiety, depression), and other symptoms (e.g., malaise and fatigue, myalgia and myositis). (**Figure 4, File S1C**).

**Figure 4:**
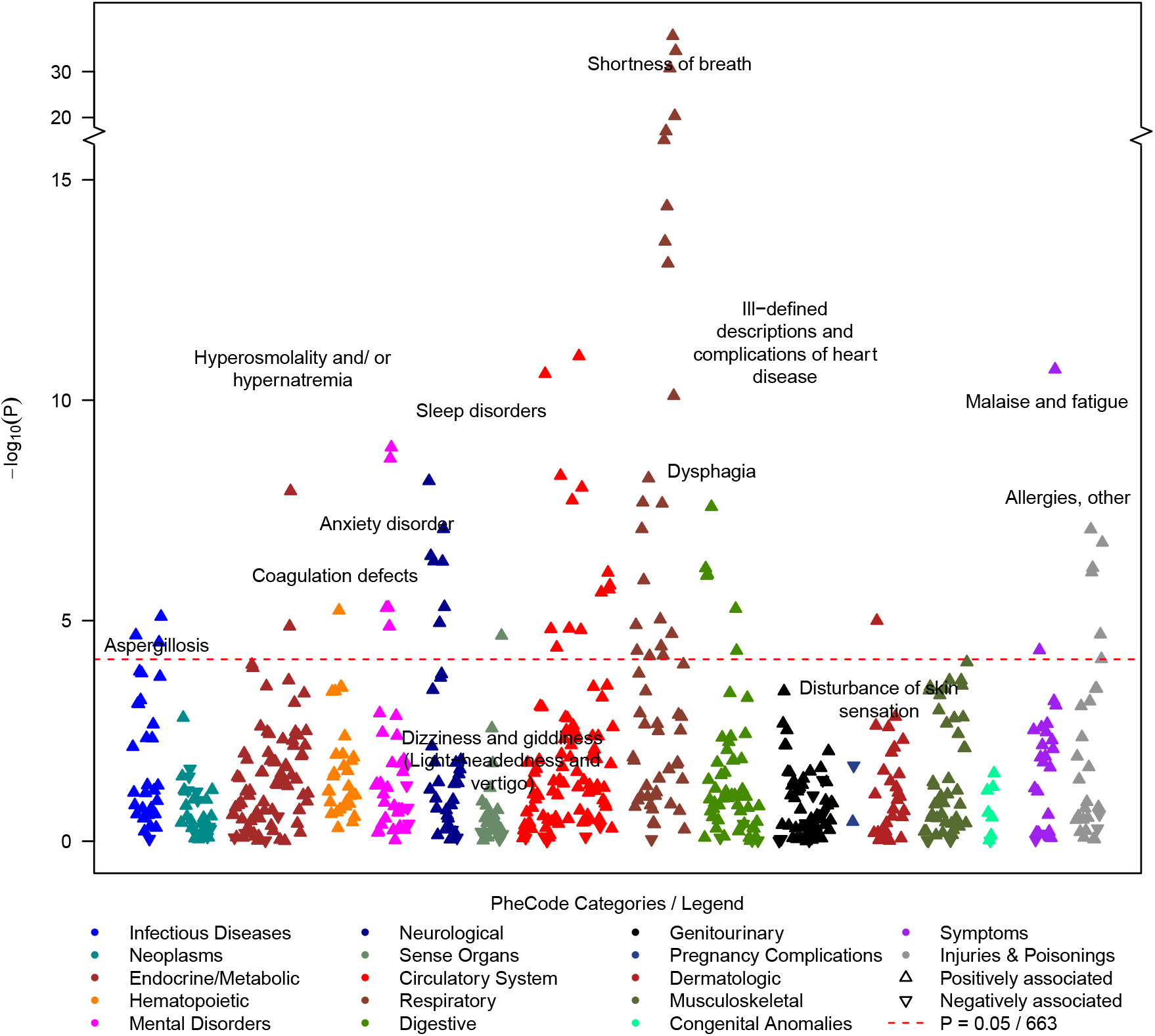
Acute-COVID-19 PheWAS on symptoms that occurred between −14 and +28 days relative to testing positive for COVID-19 (Outcome: acute-COVID-19 symptoms / PheCodes; predictor: PASC diagnosis yes/no). Among PheCodes that reached phenome-wide significance (red dashed line, P <= 0.05/663 = 7.5e-05), only the strongest association per PheCode category was labeled. The analysis was adjusted using the following covariates: age at index date, gender, race/ethnicity, Elixhauser Score AHRQ, population density (quartiles), NDI (quartiles), health care worker status, vaccination status, post-test years in EHR, and severity. Summary statistics can be found in **File S1**.

Our sensitivity analyses indicated robust associations across various settings (females only, males only, 2020 only, 2021 only, non-severe outcomes, severe outcomes) where most associations remained nominally significant in each sub-analyses or had overlapping confidence intervals in their sensitivity analyses. However, effect sizes were not as consistent (**Figures S4 A-AK, File S1G-I**). Noteworthy, the effect size for shortness of breath differed significantly between index dates in 2020 and 2021 (2020: OR = 2.20 [1.60, 2.99], P = 7.8e-7 compared to 2021: OR = 4.59 [3.62, 5.81], P = 9.37e-37; P_Difference_ = 0.000234), though they were significantly associated with PASC in both years (**Figure S4AA, File S1C&I**). Despite low numbers of individuals with severe outcomes (160 PASC cases and 150 controls), six of the 69 significantly associated phenotypes (aspergillosis, bacterial pneumonia, MRSA pneumonia, hyperosmolality and/or hypernatremia, septic shock, and voice disturbances) only had sufficient observations among the subset with severe outcomes but among the non-severe outcome subset (724 PASC cases and 6799 controls; **Table S4** and **File S1C&G**). This suggested that these phenotypes might be hospital-acquired complications. None of the 49 significantly associated phenotypes that were tested among individuals with non-severe outcomes and individuals with severe outcomes showed significant effect size differences (P_difference_ >= 0.001 [0.05/49 tests]). All phenotypes with nominal effect size differences between non-severe and severe outcomes (P_difference_ < 0.05) were all strongly and positively associated in individuals with non-severe outcomes, thus unlikely to merely represent hospital-acquired complications (**File S1G**).

### 3.5. Comparison of “pre-PASC” associated PheCode across three PheWAS

To investigate whether the associated “pre-PASC” phenotypes of the pre- and acute-COVID-19 periods (“pre-PASC” phenotypes) are causing novel PASC symptoms or if they become long-term features that manifest as PASC, we explored their frequencies and their association signals across all three PheWAS (**Figure S5**). Interestingly, almost all associated “pre-PASC” phenotypes were also significantly enriched in the post-COVID-19 PheWAS, except for “allergic reaction to food” of the pre-COVID-19 PheWAS and “candidiasis” and “inflammation and edema of the lung” in the acute-COVID-19 PheWAS. However, their ORs were all positive (**File S1–3**). Since many more acute-COVID-19 phenotypes than pre-COVID-19 phenotypes remain associated also as post-COVID-19 phenotypes, this finding suggests that some of the documented PASC diagnoses, or subtypes thereof, might represent short-term consequences of an acute infection and not necessarily PASC symptoms.

### 3.6. Developing Phenotype Risk Scores for Predicting PASC

The pre- and acute-COVID-19 PheWASs indicated pre-disposing conditions for PASC. To study whether these conditions might be helpful in predicting PASC among COVID-19 positives, we generated two PheRSs: a pre-COVID-19 PheRS “PheRS1” and an acute-COVID-19 PheRS “PheRS2”. We avoided overfitting by using PheWAS results and PheRS weights obtained from individuals with index dates in 2020 or 2021, while the evaluations were performed in individuals with index dates in 2022 (**Figure 1, Figure S2**, and **File S1J**). To limit the impact of potential hospital-acquired complications of an acute-COVID-19 infection, we excluded the six phenotypes that were only tested/observed in the individuals with severe outcomes (see “acute-COVID-19 PheWAS” above).

We found that PheRS1 and PheRS2 could discriminate cases and controls, yet only with low accuracy (AAUC < 0.7). PheRS1 performance was comparable in the complete testing data (AAUC_PheRS1_ = 0.548 [95% CI: 0.516, 0.580]) and the testing data that was reduced to PASC cases that had at least 28 days between their index date and the PASC diagnosis (AAUC_PheRS1_ = 0.555 [95% CI: 0.496, 0.612]). PheRS2 was only analyzed in the latter data (AAUC_PheRS2_ = 0.605 [95% CI: 0.549, 0.663]) but performed better than PheRS1, which was also evident from its pseudo-R^2^ which was almost 5-fold higher (0.0116 and 0.0547, respectively). A combination score further improved the discrimination of cases and controls, but its accuracy remained low (AAUC_Combined_ = 0.615 [0.561, 0.670]; **Table 2)**. We also explored if PheRSs based on additional suggestively associated PheCodes (defined as P < 1E-3) could further improve the prediction of PASC but found their individual or combined predictive ability slightly worse compared to the PheRSs that were based on phenome-wide significant hits (e.g., AAUC_Combined_ = 0.601 [0.548, 0.658]; **Table S7)**.

**Table 2:** PheRS Evaluation in the testing data (COVID-19 positive in 2022). PheRS1 was based on the significant hits of the PheWAS with the pre-COVID-19 training data (1,256 cases and 11,674 controls; COVID-19 positive in 2020/2021) while PheRS2 was based on the significant hits of the PheWAS with the acute-COVID-19 training data (874 cases and 8,144 controls; COVID-19 positive in 2020/2021 & at least 28 days between first COVID-19 and first PASC diagnosis). Underlying weights can be found in **File S2 and Table S8**.

| Predictor | Testing Data |  | AAUC <sup>a</sup><br>(95% CI) | Pseudo-R<sup>2</sup> <sup>b</sup> | Brier Score |
| --- | --- | --- | --- | --- | --- |
|  | n Cases | n Controls |  |  |  |
| PheRS1 | 349 | 3248 | 0.548 (0.516, 0.580) | n/a <sup>c</sup> | n/a <sup>c</sup> |
| PheRS1 | 123 | 1154 | 0.555 (0.496, 0.612) | 0.0116 | 0.0857 |
| PheRS2 |  |  | 0.605 (0.549, 0.663) | 0.0547 | 0.0823 |
| PheRS1 & PheRS2 |  |  | 0.615 (0.561, 0.670) | 0.0553 | 0.0824 |

<sup>a</sup> Adjusted for age at index date, gender, race/ethnicity, Elixhauser Score, population density, NDI, health care worker status, vaccination status, pre-test years in EHR, and severity
<sup>b</sup> Nagelkerke [Cragg and Uhler])
<sup>c</sup> not applicable, only useful in evaluating multiple models predicting the same outcome on the same dataset

While the use for individual-level prediction seemed very limited, we found that PheRS1 and PheRS2 could significantly enrich PASC cases in their top 10% and top 10-25% risk bins compared to the lower 50% of their distributions (**Table 3**). For example, individuals in the top 10% of PheRS1 were 2.5 times (OR = 2.48 [95% CI: 1.24, 4.97]) and in the top 10% of PheRS2 4.1 times more likely to obtain a PASC diagnosis (OR: 4.10 [2.28, 7.40]). Moreover, both PheRSs combined improved enrichment also in the top 10-25% risk bin (OR: 2.91 [1.73, 4.90]), identifying a fourth of all COVID-19 cases with substantially increased risk for PASC (**Table 3**).

**Table 3:** PheRS-based risk stratification in the testing data. Analysis is based on COVID-19 positive individuals in 2022 with at least 28 days between first COVID-19 and first PASC diagnosis; 123 cases and 1154 controls.

| PheRS | Upper Risk Bin | %Cases in Risk Bin | %Cases in Lower 50% | OR (95% CI)<sup>a</sup> | P |
| --- | --- | --- | --- | --- | --- |
| PheRS1 | 25-50% | 10.0 | 7.8 | 1.48 (0.91, 2.42) | 0.12 |
|  | 10-25% | 12.1 |  | 1.86 (1.06, 3.25) | 0.029 |
|  | >=10% | 13.6 |  | 2.48 (1.24, 4.97) | 0.011 |
| PheRS2 | 25-50% | 8.1 | 6.6 | 1.26 (0.76, 2.08) | 0.38 |
|  | 10-25% | 12.6 |  | 2.13 (1.25, 3.62) | 0.0053 |
|  | >=10% | 21.6 |  | 4.10 (2.28, 7.40) | 2.70E-06 |
| PheRS1 & PheRS2 | 25-50% | 8.3 | 6.2 | 1.36 (0.82, 2.28) | 0.23 |
|  | 10-25% | 15.2 |  | 2.91 (1.73, 4.90) | 5.80E-05 |
|  | >=10% | 19.4 |  | 3.94 (2.10, 7.42) | 2.10E-05 |

<sup>a</sup> Enrichment of PASC cases in risk bin compared to lower 50%; adjusted for age at index date, gender, race/ethnicity, Elixhauser Score, population density, NDI, health care worker status, vaccination status, pre-test years in EHR, and severity

## 4. Discussion

In this study, we used data from a relatively large cohort of COVID-19-positive individuals from MM, a single medical center. We applied a PheWAS approach across time-restricted phenomes to identify phenotypes that may predispose to PASC. We found seven phenotypes (e.g., IBS, concussion, shortness of breath) of the pre-COVID-19 period and 69 phenotypes (predominantly respiratory and circulatory symptoms) of the acute-COVID-19 period to be significantly enriched among PASC cases. Most of them were also observed enriched among PASC cases in the post-COVID19 period indicating that some of these phenotypes might have become longer-lasting or even chronic conditions. When incorporating these findings into PheRSs, we found that both the pre-COVID-19 PheRS and the acute-COVID-19 PheRS could predict PASC only with low accuracy among COVID-19-positive individuals, even when combined. However, both combined PheRSs could identify a quarter of COVID-19 positives with at least 2.9-fold increased risk of PASC.

A comparison of our findings with previous studies confirmed many pre-existing conditions that predispose to PASC. For example, in the pre-COVID-19 period PheWAS, we identified several respiratory symptoms that predisposed to PASC, including shortness of breath and other respiratory abnormalities. These findings are consistent with previous works [15, 27, 49]. The literature on IBS as a pre-disposing diagnosis for PASC seems sparse; however, there might be a connection between gut microbiota and the clinical course of COVID-19 [50] and mediation of risk factors effects for COVID-19 [51, 52]. Similarly little seems to be known of concussion as a pre-disposing diagnosis for PASC; yet, pre-existing cognitive risk factors like mild traumatic brain injury were reported as enriched among cognitive PASC cases compared to non-cognitive PASC patients [53]. Future studies are needed to substantiate our findings and investigate how pre-disposing diagnoses relate to PASC. In addition to the results from the pre-COVID-19 period conditions, our findings from the acute-COVID-19 period also accord with previous studies. Among the 69 PASC-associated phenotypes, the majority were respiratory symptoms and in line with earlier reports (e.g., cough [54, 55], dyspnea [56], respiratory insufficiency [57]). Also, the identified muscle-related symptoms, including myalgia, malaise and fatigue, were supported by previous PASC studies [58, 59]. Similar to the findings of Xie et al., we found circulatory diseases to play an essential role as a predisposing factor for PASC. While not all observed associations were previously reported, our sensitivity analyses indicated overall robustness across various settings [61, 62].

An overlap between the enriched symptoms in the three periods implies the possibility of PASC being recurring symptoms of pre-existing conditions [17]. The difference in subsiding rate between cases and controls in some symptoms (e.g., respiratory symptoms) potentially indicates the development of chronic conditions [9, 63].

There are several limitations to our analysis. First, we focused on predisposing diagnoses and performed matching, incl. on age, gender, and race/ethnicity, to adjust for potential confounding; however, these demographic characteristics were previously implicated as pre-disposing factors [64-66]. So, while matching and adjusting for these covariates might have effectively increased the power to identify pre-existing phenotypes that increase the risk for PASC, we disregarded these demographic factors as PASC predictors. Future studies are needed to evaluate the combined contributions of these variables in more comprehensive prediction models. Second, although a clinical diagnosis of PASC was used, many reported symptoms are vague, unspecific, and subtle [67], and awareness about PASC only recently increased. This might lead to an underdiagnosis of PASC [68, 69]. For example, we only observed 2.7% PASC-diagnosed patients in our COVID-19 positive cohort, which is far lower than PASC studies from the US, which estimated a prevalence between 19% and 35% [70]. As a result, our predictions of PASC might be overly conservative. The available diagnosis codes for PASC lacked specificity to stratify PASC cases into PASC subtypes reliably. Future studies that incorporate natural language processing of clinical notes and that have larger sample sizes will likely improve the identification of PASC cases and subtypes [71]. Third, the analysis was restricted to the COVID-19-positive individuals who were also seen at MM during the pre-COVID-19 and the post-COVID-19 periods; due to this selection bias, both cases and controls might be less healthy and older compared to randomly chosen COVID-19-positive individuals [72].

Moreover, it has been reported that around 15% - 40% of the confirmed COVID-19 population were asymptomatic [73, 74]. Using data from a health system caused our cohort to be enriched for symptomatic COVID-19 patients, while asymptomatic COVID-19 cases may be underrepresented. Such biases and omissions might limit the generalizability to the overall population. Although this study included a large size of COVID-19 patients, attention might be given to expanding and diversifying the collection and analysis of data.

Our study used a clinical definition of PASC. In addition to the commonly used ICD code U09.9 (“Post COVID-19 condition, unspecified”) or B94.8 (“Sequelae of other specified infectious and parasitic diseases”), we applied the information from the EHR internal problem list database (PSL, **Table S1**) to categorize PASC patients, which enabled us to collect patients whose diagnosis were recorded even before official ICD-10 recommendations/codes became available. The post-COVID-19 period PheWAS validated our PASC definition in that we enriched diagnoses consistent with subtypes of PASC that were previously reported (e.g., shortness of breath, neurological disorders, malaise, fatigue and dysphagia) [3, 71, 75]. Furthermore, given the benefit of rich retrospective EHR data, we could adjust for essential confounders in our models, including race, Elixhauser comorbidity score, vaccination status, etc., that might have affected PASC outcomes. We expect that our approach and the resulting prediction models will improve over time with increasing sample sizes and, by doing so, will likely facilitate earlier detection of PASC cases or improve risk stratification. Furthermore, a better characterization of PASC mechanisms might inform on distinct PASC forms that differ in their profiles of pre-existing conditions.

## 5. Conclusions

PASC represents a worldwide public health challenge affecting millions of people. While effective therapies for PASC are still in development [76-79], prediction and risk models can help to more reliably identify individuals at increased risk for PASC and its subcategories and potentially inform preventive or therapeutic efforts.

The present research aimed to identify PASC pre-disposing diagnoses from the pre- and the acute-COVID-19 medical phenomes and to explore them as predictors for PASC. We identified known and potentially novel associations across various disease categories in both phenomes. These phenotypes, when aggregated into PheRSs, have predictive properties for PASC, especially when considered for risk stratification approaches. Future studies might consider applying more complex non-linear models like machine learning to improve prediction models. The next opportunity will be to incorporate additional, more complex data like laboratory measurements or medication data into such prediction models, as they have proven relevant for PASC but have yet to be fully investigated [2, 80, 81]. The presented PheRS framework can also be adapted to explore alternative outcomes like survival and, by doing so, offer comprehensive insights into the long-term consequences of COVID-19.

## Data Availability

Electronic Health Record Data cannot be shared publicly as a result of patient confidentiality. Summary statistics are contained in the supplementary material of the manuscript.

## Acknowledgement

The authors acknowledge Precision Health at the University of Michigan, and the University of Michigan Medical School Data Office for Clinical and Translational Research for providing data storage, management, processing, and distribution services. This work does not represent the views of the US Government or the Department of Veterans Affairs.

## CRediT author statement

**Lars G. Fritsche:** Conceptualization, Methodology, Formal analysis, Investigation, Data Curation, Writing - Original Draft, Writing - Review & Editing, Visualization, Funding acquisition. **Weijia Jin:** Writing - Original Draft, Writing - Review & Editing, Visualization. **Andrew J. Admon:** Writing - Review & Editing. **Bhramar Mukherjee:** Conceptualization, Methodology, Writing - Original Draft, Writing - Review & Editing, Supervision, Funding acquisition.

## References

[1] Microsoft Corporation. Bing COVID-19 Tracker. 2022.

[2] Al-Aly Z, Xie Y, Bowe B. High-dimensional characterization of post-acute sequelae of COVID-19. Nature. 2021;594:259–64.

[3] Chen C, Haupert SR, Zimmermann L, Shi X, Fritsche LG, Mukherjee B. Global Prevalence of Post COVID-19 Condition or Long COVID: A Meta-Analysis and Systematic Review. J Infect Dis. 2022.

[4] Lopez-Leon S, Wegman-Ostrosky T, Ayuzo Del Valle NC, Perelman C, Sepulveda R, Rebolledo PA, et al. Long-COVID in children and adolescents: a systematic review and meta-analyses. Sci Rep. 2022;12:9950.

[5] Centers for Disease Control and Prevention. Post-COVID Conditions: Information for Healthcare Providers. Avaliable: https://www.cdc.gov/coronavirus/2019-ncov/hcp/clinical-care/post-covid-index.html. https://www.cdc.gov/coronavirus/2019-ncov/hcp/clinical-care/post-covid-index.html2021.

[6] Centers for Disease Control and Prevention. Public Health Recommendations. Avaliable: https://www.cdc.gov/coronavirus/2019-ncov/hcp/clinical-care/post-covid-public-health-recs.html. https://www.cdc.gov/coronavirus/2019-ncov/hcp/clinical-care/post-covid-public-health-recs.html2021.

[7] Centers for Disease Control and Prevention. Long COVID or Post-COVID Conditions. Avaliable: https://www.cdc.gov/coronavirus/2019-ncov/long-term-effects/index.html. https://www.cdc.gov/coronavirus/2019-ncov/hcp/clinical-care/post-covid-conditions.html2021.

[8] Nalbandian A, Sehgal K, Gupta A, Madhavan MV, McGroder C, Stevens JS, et al. Post-acute COVID-19 syndrome. Nat Med. 2021;27:601–15.

[9] Baig AM. Chronic COVID syndrome: Need for an appropriate medical terminology for long-COVID and COVID long-haulers. J Med Virol. 2021;93:2555–6.

[10] Nath A. Long-Haul COVID. Neurology. 2020;95:559–60.

[11] Aiyegbusi OL, Hughes SE, Turner G, Rivera SC, McMullan C, Chandan JS, et al. Symptoms, complications and management of long COVID: a review. J R Soc Med. 2021;114:428–42.

[12] Kamal M, Abo Omirah M, Hussein A, Saeed H. Assessment and characterisation of post-COVID-19 manifestations. Int J Clin Pract. 2021;75:e13746.

[13] Huang C, Huang L, Wang Y, Li X, Ren L, Gu X, et al. 6-month consequences of COVID-19 in patients discharged from hospital: a cohort study. Lancet. 2021;397:220–32.

[14] Chippa V, Aleem A, Anjum F. Post Acute Coronavirus (COVID-19) Syndrome. StatPearls. Treasure Island (FL): StatPearls Publishing Copyright © 2022, StatPearls Publishing LLC.; 2022.

[15] Daher A, Balfanz P, Cornelissen C, Müller A, Bergs I, Marx N, et al. Follow up of patients with severe coronavirus disease 2019 (COVID-19): Pulmonary and extrapulmonary disease sequelae. Respiratory medicine. 2020;174:106197.

[16] Stefanou MI, Palaiodimou L, Bakola E, Smyrnis N, Papadopoulou M, Paraskevas GP, et al. Neurological manifestations of long-COVID syndrome: a narrative review. Ther Adv Chronic Dis. 2022;13:20406223221076890.

[17] Davis HE, Assaf GS, McCorkell L, Wei H, Low RJ, Re’em Y, et al. Characterizing long COVID in an international cohort: 7 months of symptoms and their impact. EClinicalMedicine. 2021;38:101019.

[18] Taquet M, Sillett R, Zhu L, Mendel J, Camplisson I, Dercon Q, et al. Neurological and psychiatric risk trajectories after SARS-CoV-2 infection: an analysis of 2-year retrospective cohort studies including 1 284 437 patients. Lancet Psychiatry. 2022.

[19] Premraj L, Kannapadi NV, Briggs J, Seal SM, Battaglini D, Fanning J, et al. Mid and long-term neurological and neuropsychiatric manifestations of post-COVID-19 syndrome: A meta-analysis. J Neurol Sci. 2022;434:120162.

[20] Wang W, Wang CY, Wang SI, Wei JC. Long-term cardiovascular outcomes in COVID-19 survivors among non-vaccinated population: A retrospective cohort study from the TriNetX US collaborative networks. EClinicalMedicine. 2022;53:101619.

[21] Xu E, Xie Y, Al-Aly Z. Long-term neurologic outcomes of COVID-19. Nat Med. 2022.

[22] Ayoubkhani D, Bermingham C, Pouwels KB, Glickman M, Nafilyan V, Zaccardi F, et al. Trajectory of long covid symptoms after covid-19 vaccination: community based cohort study. Bmj. 2022;377:e069676.

[23] Al-Aly Z, Bowe B, Xie Y. Long COVID after breakthrough SARS-CoV-2 infection. Nature Medicine. 2022.

[24] Bai F, Tomasoni D, Falcinella C, Barbanotti D, Castoldi R, Mulè G, et al. Female gender is associated with long COVID syndrome: a prospective cohort study. Clin Microbiol Infect. 2022;28:611.e9-.e16.

[25] Antonelli M, Pujol JC, Spector TD, Ourselin S, Steves CJ. Risk of long COVID associated with delta versus omicron variants of SARS-CoV-2. Lancet. 2022;399:2263–4.

[26] Yoo SM, Liu TC, Motwani Y, Sim MS, Viswanathan N, Samras N, et al. Factors Associated with Post-Acute Sequelae of SARS-CoV-2 (PASC) After Diagnosis of Symptomatic COVID-19 in the Inpatient and Outpatient Setting in a Diverse Cohort. J Gen Intern Med. 2022;37:1988–95.

[27] Sudre CH, Murray B, Varsavsky T, Graham MS, Penfold RS, Bowyer RC, et al. Attributes and predictors of long COVID. Nat Med. 2021;27:626–31.

[28] Cervia C, Zurbuchen Y, Taeschler P, Ballouz T, Menges D, Hasler S, et al. Immunoglobulin signature predicts risk of post-acute COVID-19 syndrome. Nat Commun. 2022;13:446.

[29] Pfaff ER, Girvin AT, Bennett TD, Bhatia A, Brooks IM, Deer RR, et al. Identifying who has long COVID in the USA: a machine learning approach using N3C data. Lancet Digit Health. 2022;4:e532–41.

[30] Su Y, Yuan D, Chen DG, Ng RH, Wang K, Choi J, et al. Multiple early factors anticipate post-acute COVID-19 sequelae. Cell. 2022;185:881–95 e20.

[31] Salvatore M, Beesley LJ, Fritsche LG, Hanauer D, Shi X, Mondul AM, et al. Phenotype risk scores (PheRS) for pancreatic cancer using time-stamped electronic health record data: Discovery and validation in two large biobanks. J Biomed Inform. 2021;113:103652.

[32] Salvatore M, Gu T, Mack JA, Prabhu Sankar S, Patil S, Valley TS, et al. A Phenome-Wide Association Study (PheWAS) of COVID-19 Outcomes by Race Using the Electronic Health Records Data in Michigan Medicine. J Clin Med. 2021;10.

[33] Estiri H, Strasser ZH, Brat GA, Semenov YR, Consortium for Characterization of C-bEHR, Patel CJ, et al. Evolving phenotypes of non-hospitalized patients that indicate long COVID. BMC Med. 2021;19:249.

[34] National Center for Immunization and Respiratory Diseases (NCIRD); Division of Viral Diseases. Evaluating and Caring for Patients with Post-COVID Conditions: Interim Guidance. 2021.

[35] Clarke P, Melendez R. National Neighborhood Data Archive (NaNDA): Neighborhood Socioeconomic and Demographic Characteristics by Tract, United States, 2000-2010. In: National Neighborhood Data Archive (NaNDA), editor. openICPSR-111107, nanda_ses2000-2010_01P.* ed 2019.

[36] Melendez R, Clarke P, Khan A, Gomez-Lopez I, Li M, Chenoweth M. National Neighborhood Data Archive (NaNDA): Socioeconomic Status and Demographic Characteristics of ZIP Code Tabulation Areas, United States, 2008-2017. ICPSR - Interuniversity Consortium for Political and Social Research. 2020.

[37] Gasparini A. comorbidity: An R package for computing comorbidity scores. Journal of Open Source Software. 2018;3:648.

[38] Elixhauser A, Steiner C, Harris DR, Coffey RM. Comorbidity measures for use with administrative data. Med Care. 1998;36:8–27.

[39] Wu P, Gifford A, Meng X, Li X, Campbell H, Varley T, et al. Mapping ICD-10 and ICD-10-CM Codes to Phecodes: Workflow Development and Initial Evaluation. JMIR Med Inform. 2019;7:e14325.

[40] Carroll RJ, Bastarache L, Denny JC. R PheWAS: data analysis and plotting tools for phenome-wide association studies in the R environment. Bioinformatics. 2014;30:2375–6.

[41] Ho DE, Imai K, King G, Stuart EA. MatchIt: Nonparametric Preprocessing for Parametric Causal Inference. J Stat Softw. 2011;42:1–28.

[42] Friedman JH, Hastie T, Tibshirani R. Regularization Paths for Generalized Linear Models via Coordinate Descent. J Stat Softw. 2010;33:1 – 22.

[43] Hoerl AE, Kennard RW. Ridge Regression: Biased Estimation for Nonorthogonal Problems. Technometrics. 1970;12:55–67.

[44] Mangiafico S. rcompanion: Functions to Support Extension Education Program Evaluation. 2021.

[45] Signorell A. {DescTools}: Tools for Descriptive Statistics. 2021.

[46] Rodríguez-Álvarez MX, Iácio V. {ROCnReg}: An {R} Package for Receiver Operating Characteristic Curve Inference With and Without Covariates. The R Journal. 2021;13:525–55.

[47] Kosmidis I. {brglm2}: Bias Reduction in Generalized Linear Models. 2021.

[48] R Core Team. R: A Language and Environment for Statistical Computing. Vienna, Austria: R Foundation for Statistical Computing; 2022.

[49] Osmanov IM, Spiridonova E, Bobkova P, Gamirova A, Shikhaleva A, Andreeva M, et al. Risk factors for post-COVID-19 condition in previously hospitalised children using the ISARIC Global follow-up protocol: a prospective cohort study. Eur Respir J. 2022;59.

[50] Vodnar DC, Mitrea L, Teleky BE, Szabo K, Calinoiu LF, Nemes SA, et al. Coronavirus Disease (COVID-19) Caused by (SARS-CoV-2) Infections: A Real Challenge for Human Gut Microbiota. Front Cell Infect Microbiol. 2020;10:575559.

[51] Chen J, Hall S, Vitetta L. Altered gut microbial metabolites could mediate the effects of risk factors in Covid-19. Rev Med Virol. 2021;31:1–13.

[52] Chen J, Vitetta L. Gut-brain axis in the neurological comorbidity of COVID-19. Brain Commun. 2021;3:fcab118.

[53] Apple AC, Oddi A, Peluso MJ, Asken BM, Henrich TJ, Kelly JD, et al. Risk factors and abnormal cerebrospinal fluid associate with cognitive symptoms after mild COVID-19. Ann Clin Transl Neurol. 2022;9:221–6.

[54] Jennings G, Monaghan A, Xue F, Mockler D, Romero-Ortuño R. A Systematic Review of Persistent Symptoms and Residual Abnormal Functioning following Acute COVID-19: Ongoing Symptomatic Phase vs. Post-COVID-19 Syndrome. J Clin Med. 2021;10.

[55] Kang YR, Oh JY, Lee JH, Small PM, Chung KF, Song WJ. Long-COVID severe refractory cough: discussion of a case with 6-week longitudinal cough characterization. Asia Pac Allergy. 2022;12:e19.

[56] Fernández-de-las-Peñas C, Pellicer-Valero OJ, Navarro-Pardo E, Palacios-Ceña D, Florencio LL, Guijarro C, et al. Symptoms Experienced at the Acute Phase of SARS-CoV-2 Infection as Risk Factor of Long-term Post-COVID Symptoms: The LONG-COVID-EXP-CM Multicenter Study. International Journal of Infectious Diseases. 2022;116:241–4.

[57] Cabrera Martimbianco AL, Pacheco RL, Bagattini Â M, Riera R. Frequency, signs and symptoms, and criteria adopted for long COVID-19: A systematic review. Int J Clin Pract. 2021;75:e14357.

[58] Petersen MS, Kristiansen MF, Hanusson KD, Danielsen ME, B ÁS, Gaini S, et al. Long COVID in the Faroe Islands: A Longitudinal Study Among Nonhospitalized Patients. Clin Infect Dis. 2021;73:e4058–e63.

[59] Soares MN, Eggelbusch M, Naddaf E, Gerrits KHL, van der Schaaf M, van den Borst B, et al. Skeletal muscle alterations in patients with acute Covid-19 and post-acute sequelae of Covid-19. J Cachexia Sarcopenia Muscle. 2022;13:11–22.

[60] Xie Y, Xu E, Bowe B, Al-Aly Z. Long-term cardiovascular outcomes of COVID-19. Nat Med. 2022;28:583–90.

[61] Thabane L, Mbuagbaw L, Zhang S, Samaan Z, Marcucci M, Ye C, et al. A tutorial on sensitivity analyses in clinical trials: the what, why, when and how. BMC Med Res Methodol. 2013;13:92.

[62] Borgonovo E, Plischke E. Sensitivity analysis: A review of recent advances. European Journal of Operational Research. 2016;248:869–87.

[63] Bell ML, Catalfamo CJ, Farland LV, Ernst KC, Jacobs ET, Klimentidis YC, et al. Post-acute sequelae of COVID-19 in a non-hospitalized cohort: Results from the Arizona CoVHORT. PLoS One. 2021;16:e0254347.

[64] Thompson EJ, Williams DM, Walker AJ, Mitchell RE, Niedzwiedz CL, Yang TC, et al. Long COVID burden and risk factors in 10 UK longitudinal studies and electronic health records. Nat Commun. 2022;13:3528.

[65] Whitaker M, Elliott J, Chadeau-Hyam M, Riley S, Darzi A, Cooke G, et al. Persistent COVID-19 symptoms in a community study of 606,434 people in England. Nat Commun. 2022;13:1957.

[66] Clinical characteristics with inflammation profiling of long COVID and association with 1-year recovery following hospitalisation in the UK: a prospective observational study. Lancet Respir Med. 2022.

[67] Greenhalgh T, Knight M, A’Court C, Buxton M, Husain L. Management of post-acute covid-19 in primary care. Bmj. 2020;370:m3026.

[68] Brackel CLH, Lap CR, Buddingh EP, van Houten MA, van der Sande L, Langereis EJ, et al. Pediatric long-COVID: An overlooked phenomenon? Pediatr Pulmonol. 2021;56:2495–502.

[69] Parkin A, Davison J, Tarrant R, Ross D, Halpin S, Simms A, et al. A Multidisciplinary NHS COVID-19 Service to Manage Post-COVID-19 Syndrome in the Community. J Prim Care Community Health. 2021;12:21501327211010994.

[70] National Center for Health Statistics. Long COVID Household Pulse Survey. Avaliable: https://www.cdc.gov/nchs/covid19/pulse/long-covid.htm.

[71] Wang L, Foer D, MacPhaul E, Lo YC, Bates DW, Zhou L. PASCLex: A comprehensive post-acute sequelae of COVID-19 (PASC) symptom lexicon derived from electronic health record clinical notes. J Biomed Inform. 2022;125:103951.

[72] Tripepi G, Jager KJ, Dekker FW, Zoccali C. Selection Bias and Information Bias in Clinical Research. Nephron Clinical Practice. 2010;115:c94–c9.

[73] Ma Q, Liu J, Liu Q, Kang L, Liu R, Jing W, et al. Global Percentage of Asymptomatic SARS-CoV-2 Infections Among the Tested Population and Individuals With Confirmed COVID-19 Diagnosis: A Systematic Review and Meta-analysis. JAMA Netw Open. 2021;4:e2137257.

[74] He J, Guo Y, Mao R, Zhang J. Proportion of asymptomatic coronavirus disease 2019: A systematic review and meta-analysis. J Med Virol. 2021;93:820–30.

[75] Xie Y, Bowe B, Al-Aly Z. Burdens of post-acute sequelae of COVID-19 by severity of acute infection, demographics and health status. Nat Commun. 2021;12:6571.

[76] Gluckman TJ, Bhave NM, Allen LA, Chung EH, Spatz ES, Ammirati E, et al. 2022 ACC Expert Consensus Decision Pathway on Cardiovascular Sequelae of COVID-19 in Adults: Myocarditis and Other Myocardial Involvement, Post-Acute Sequelae of SARS-CoV-2 Infection, and Return to Play: A Report of the American College of Cardiology Solution Set Oversight Committee. J Am Coll Cardiol. 2022;79:1717–56.

[77] Kell DB, Laubscher GJ, Pretorius E. A central role for amyloid fibrin microclots in long COVID/PASC: origins and therapeutic implications. Biochem J. 2022;479:537–59.

[78] Parker AM, Brigham E, Connolly B, McPeake J, Agranovich AV, Kenes MT, et al. Addressing the post-acute sequelae of SARS-CoV-2 infection: a multidisciplinary model of care. Lancet Respir Med. 2021;9:1328–41.

[79] Centers for Disease Control and Prevention. Caring for People with Post-COVID Conditions. Avaliable: https://www.cdc.gov/coronavirus/2019-ncov/long-term-effects/care-post-covid.html. 2022.

[80] Peluso MJ, Thomas IJ, Munter SE, Deeks SG, Henrich TJ. Lack of Antinuclear Antibodies in Convalescent Coronavirus Disease 2019 Patients With Persistent Symptoms. Clin Infect Dis. 2022;74:2083–4.

[81] Groff D, Sun A, Ssentongo AE, Ba DM, Parsons N, Poudel GR, et al. Short-term and Long-term Rates of Postacute Sequelae of SARS-CoV-2 Infection: A Systematic Review. JAMA Netw Open. 2021;4:e2128568.

