## Supplementary material for "Characterizing and Predicting Post-Acute Sequelae of SARS CoV-2 infection (PASC) in a Large Academic Medical Center in the US": Text S1, Figure S1-S5, Table S1-S8

**Text S1**

Neighborhood Disadvantage Index (NDI)
The Neighborhood Disadvantage Index (NDI) without the proportion of Black includes four census indicators (proportion of female-headed families with children, the proportion of households with public assistance income or food stamps; the proportion of families with income below the federal poverty level; the proportion of population age 16+ unemployed). We did not include measures of racial distribution within this index.

Vaccination Status

We created a covariate to capture the vaccination status at the index date coded as “unvaccinated”, “after 1. vaccination”, “after full vaccination” and “after booster” using records of vaccinations for patients who received a vaccination at MM or who have a recorded vaccination record in the Michigan Care Improvement Registry (MCIR). Michigan’s immunization providers are required to report COVID vaccination to MCIR within 24 hours of administration, meaning the EHR vaccination record should be nearly complete. Among the matched case-control cohort, 11,925 individuals had at the date of their first positive test or COVID-19 diagnosis no documented vaccination and thus were considered unvaccinated. It is possible although unlikely that they may have been vaccinated elsewhere and these records were not available. A total of 7,004 individuals had at least one documented dose of a COVID-19 vaccine. According to FDA’s vaccination guideline [1], we categorized 6,000 individuals as fully vaccinated in the primary series, meaning documentation of two doses of Moderna or Pfizer-BioNTech vaccine, or a single dose of Janssen vaccine at least 21 days before the corresponding test date [2-4]. A subset of 1,646 of the fully vaccinated patients was further classified as being boosted, i.e., they received at least one additional vaccination at least 21 days after completing the primary series. The remaining 1,004 vaccinated patients who did not complete the primary series were considered “partially vaccinated”.

COVID-19 Severity
The covariate for COVID-19-related outcome severity was dichotomized as “severe”, i.e., either hospitalization or intensive care unit (ICU) admission within one month after a positive SARS-CoV-2 RT-PCR test result or COVID-19 diagnosis, or death within two months after a positive RT-PCR test or COVID-19 diagnosis. Data on hospitalizations, ICU admissions, and death were obtained from Michigan Medicine’s EHR databases as well as the Michigan Death Registry. The remaining individuals were considered “non-severe” COVID-19-related outcomes and included non-hospitalized, symptomatic, or asymptomatic COVID-19 cases.

Elixhauser comorbidity score
The Elixhauser comorbidity score developed by the Agency for Healthcare Research and Quality (AHRQ) was calculated to comprehensively characterize patients’ pre-existing comorbidity conditions using ICD9 and ICD10 codes and the R package ‘comorbidity’ [5, 6].

Healthcare worker (HCW) status
Healthcare worker (HCW) status was defined based on documented participation in an HCW survey or a SARS-CoV-2 PCR test order for HCW.

**Text S1 References**

1. Centers for Disease Control and Prevention. Use of COVID-19 Vaccines in the United States. Available at: <https://www.cdc.gov/vaccines/covid-19/clinical-considerations/covid-19-vaccines-us.html#primary-series>. Accessed 2022/03/19 2022.

2. Baden LR, El Sahly HM, Essink B, et al. Efficacy and Safety of the mRNA-1273 SARS-CoV-2 Vaccine. N Engl J Med **2021**; 384:403-16.

3. Polack FP, Thomas SJ, Kitchin N, et al. Safety and Efficacy of the BNT162b2 mRNA Covid-19 Vaccine. N Engl J Med **2020**; 383:2603-15.

4. Sadoff J, Gray G, Vandebosch A, et al. Safety and Efficacy of Single-Dose Ad26.COV2.S Vaccine against Covid-19. N Engl J Med **2021**; 384:2187-201.

5. Gasparini A. comorbidity: An R package for computing comorbidity scores. Journal of Open Source Software **2018**; 3:648.

6. Elixhauser A, Steiner C, Harris DR, Coffey RM. Comorbidity measures for use with administrative data. Med Care **1998**; 36:8-27.

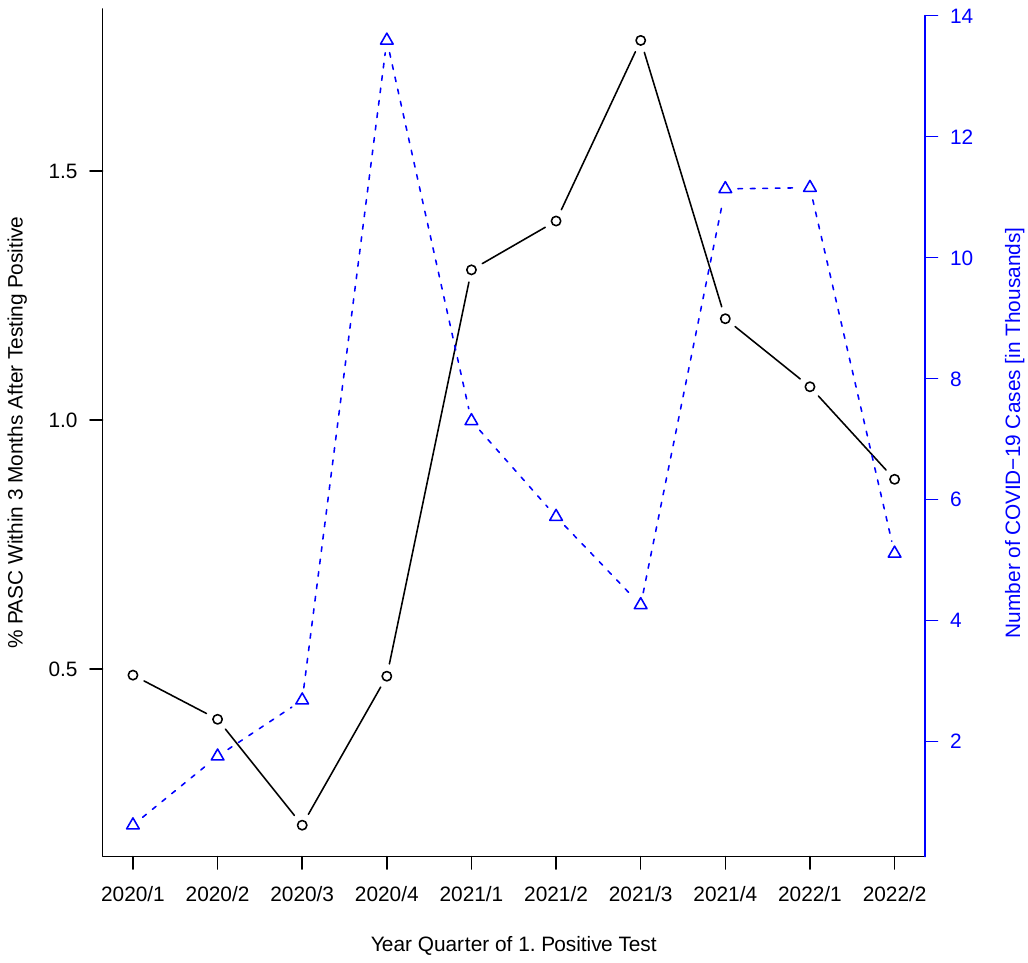

**Figure S1**. The proportion of clinically documented PASC within 3 months of testing positive and the number of total unmatched COVID-19-positive individuals by year quarter when they were tested positive/diagnosed for COVID-19 for the first time.

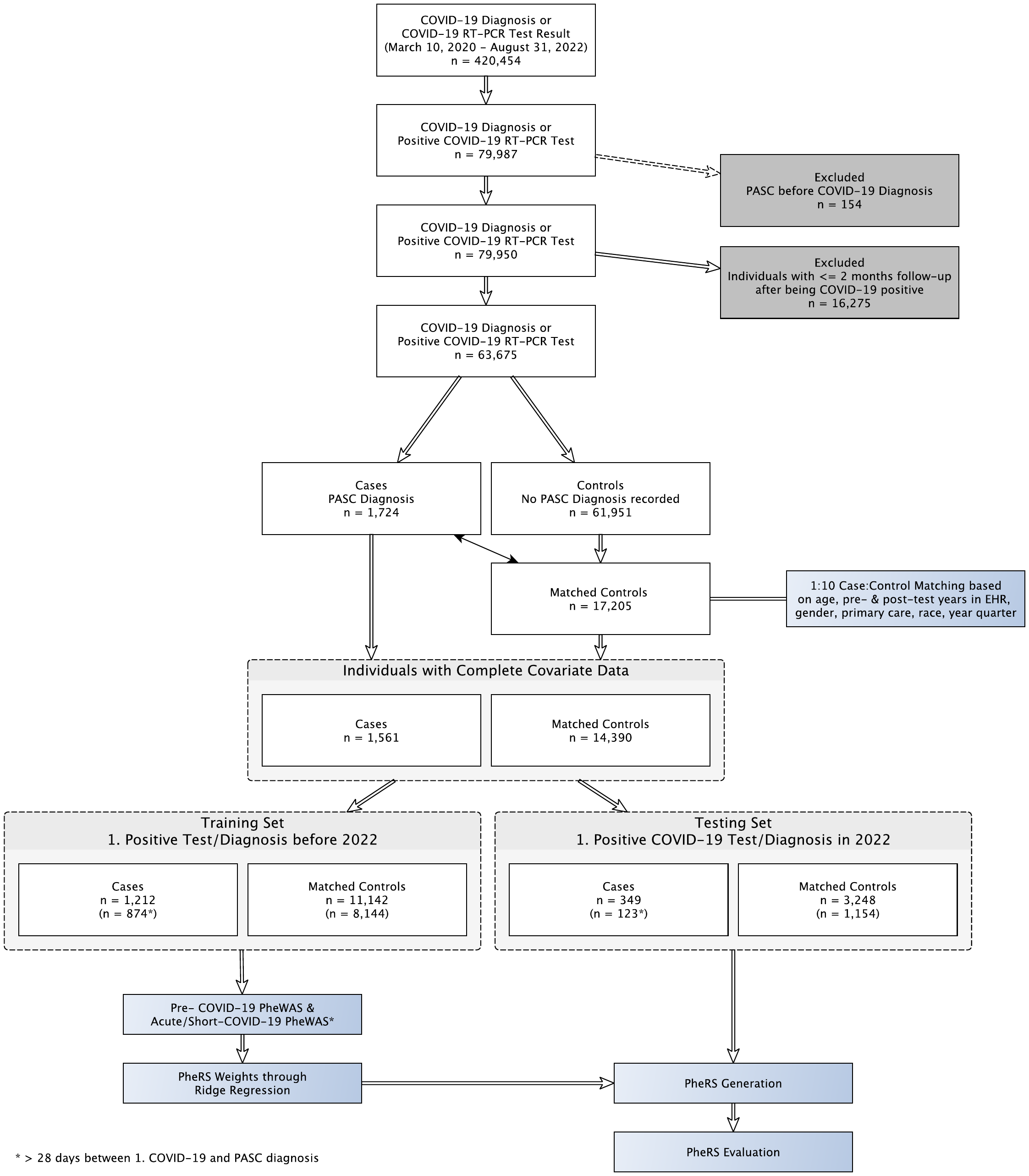

**Figure S2**: Overview flowchart showing the sample filtering and analysis setup

**Figures S3 A-G**. Forest plots of the PreCOVID-19 Sensitivity analyses. Effect sizes and PheCode frequencies in cases and controls of significantly associated PheCodes are shown. Pre-COVID-19: main analysis, only males, only females, COVID-19 positive in 2020, COVID-19 positive in 2021, non-severe COVID-19 outcome, severe COVID-19 outcome, max 2 years before COVID-19 diagnosis, and before 2020. Of significantly associated parent/child PheCodes only the phecode with the stronger association signal is shown. Sample sizes of each analysis can be found in **Table S5**.

| A | 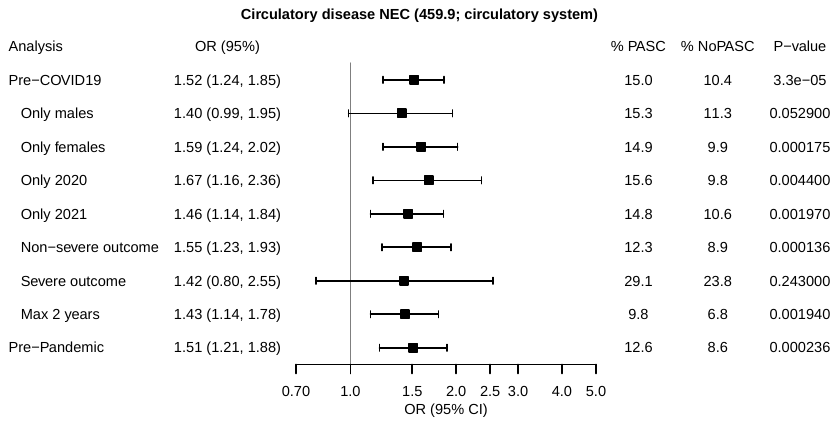 |
| --- | --- |
| B | 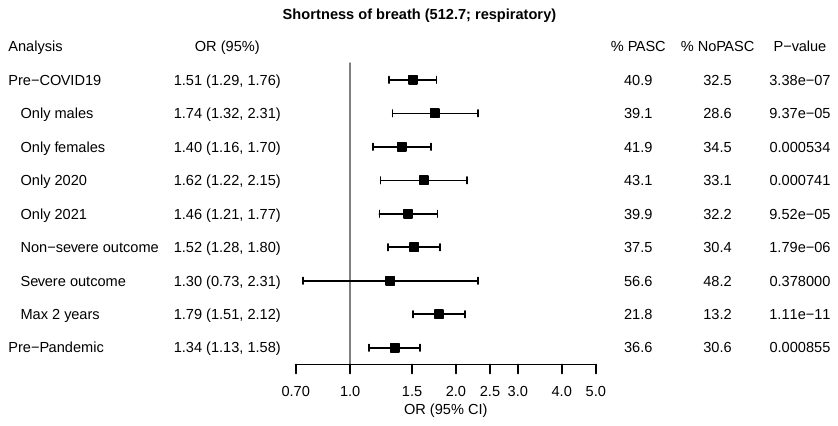 |
| C | 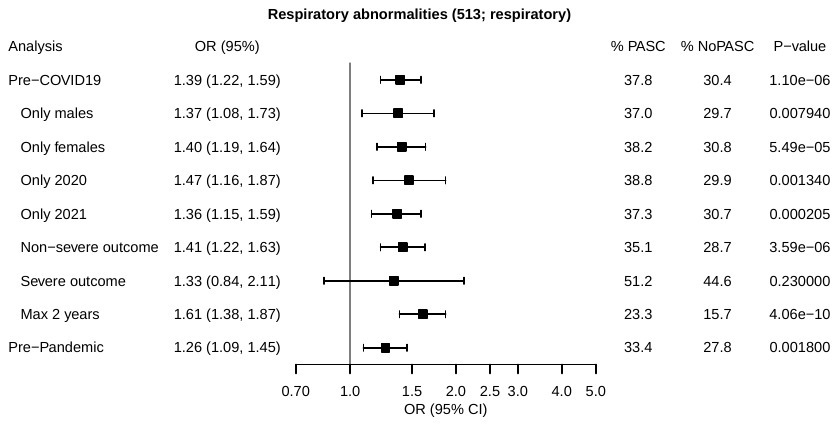 |
| D | 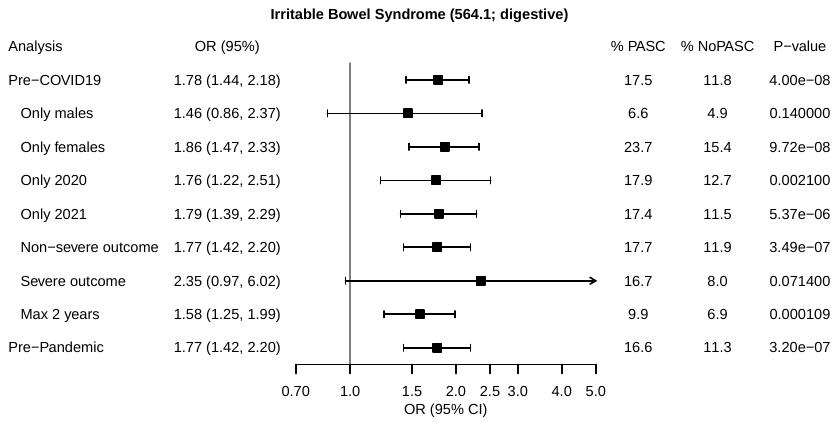 |
| E | 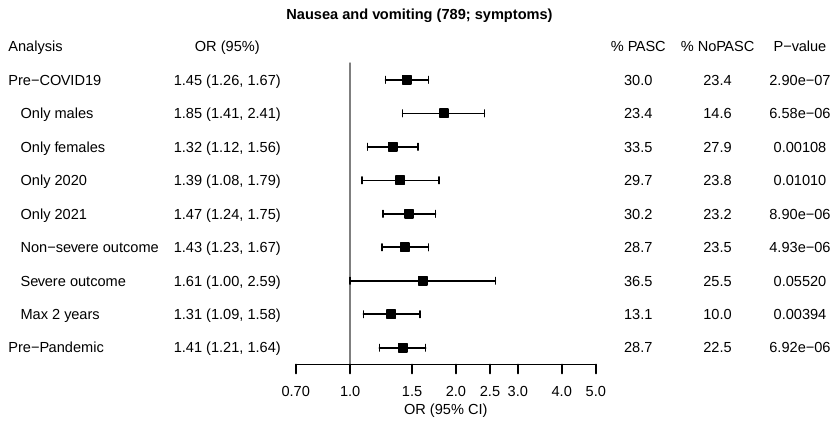 |
| F | 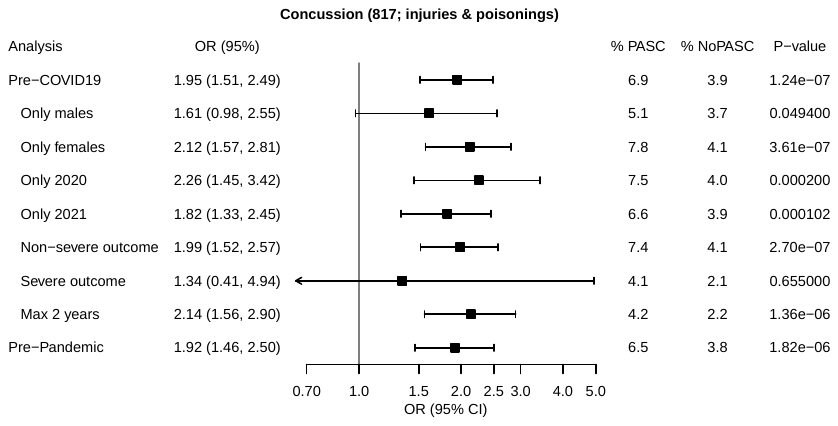 |
| G | 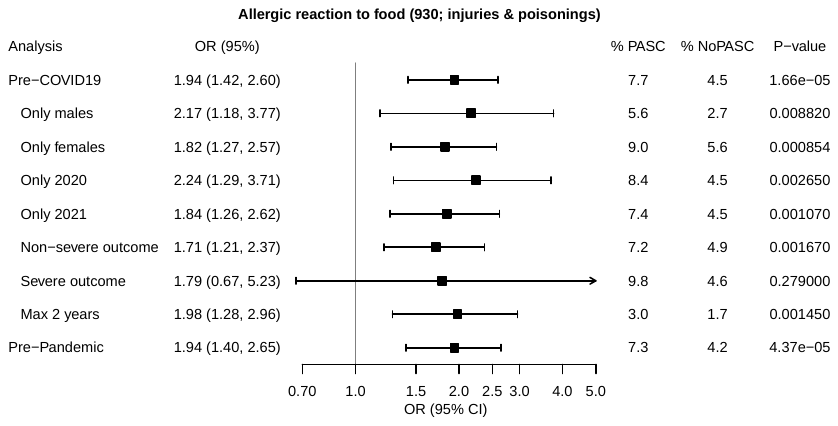 |

**Figures S4 A-Z, AA-AK**. Forest plots of the Acute & short COVID-19 Sensitivity analyses. Effect sizes and phecode frequencies in cases (PASC) and controls (NoPASC) of significantly associated PheCodes are shown. Acute & short COVID-19: main analysis, only males, only females, COVID-19 positive in 2020, COVID-19 positive in 2021, non-severe COVID-19 outcome only, and severe COVID-19 outcome. Of significantly associated parent/child phecodes only the phecode with the stronger association signal is shown. Sample sizes of each analysis can be found in **Table S5**.

| A | 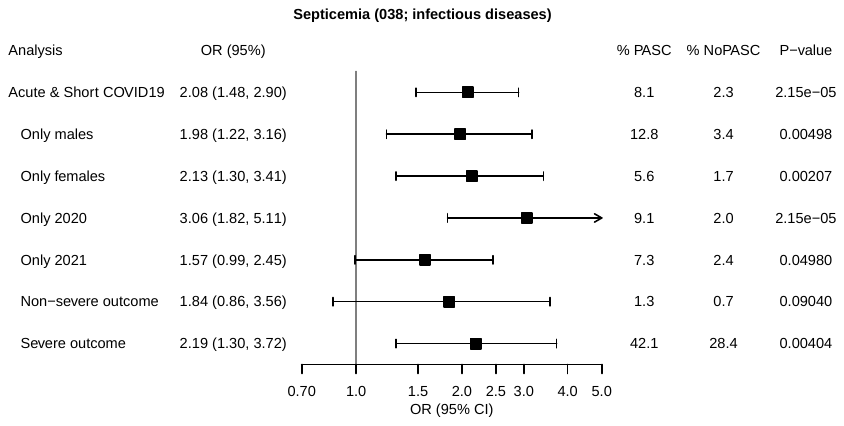 |
| --- | --- |
| B | 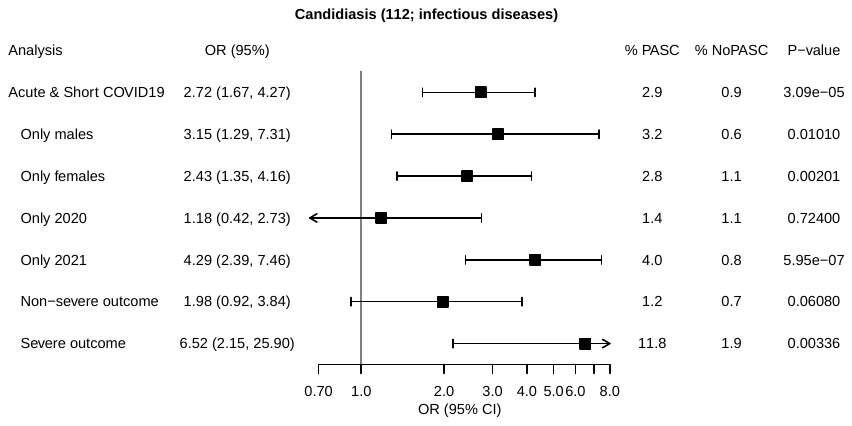 |
| C | 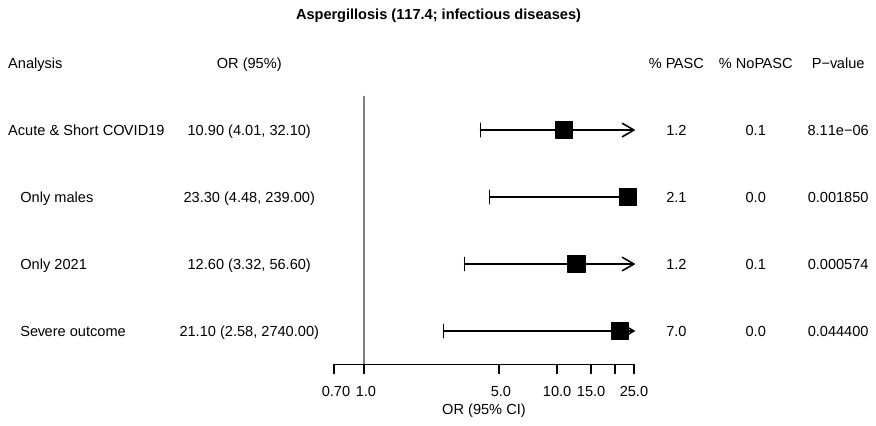 |
| D | 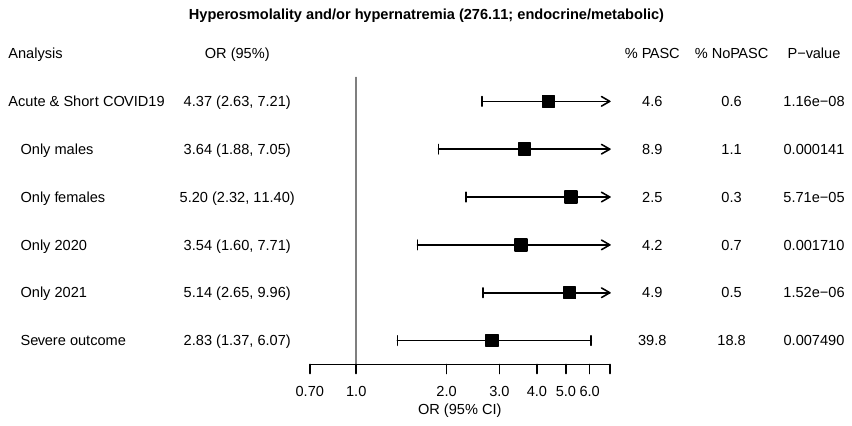 |
| E | 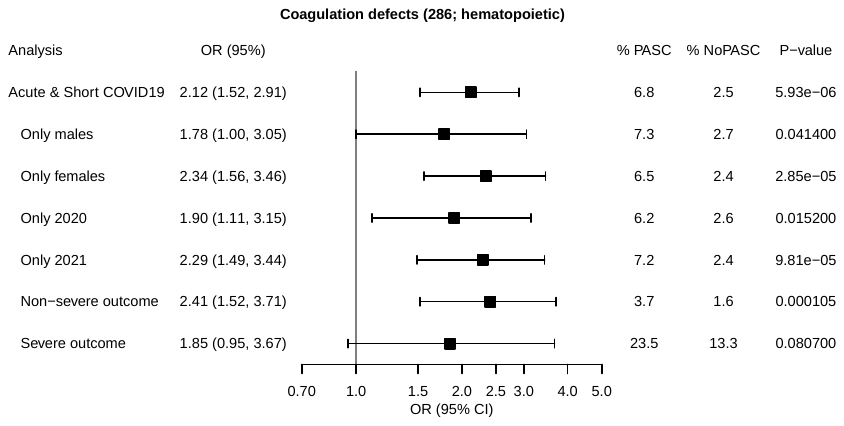 |
| F | 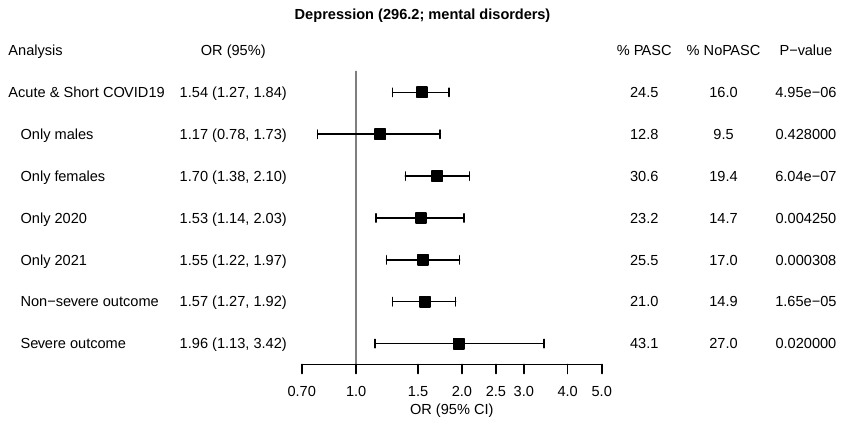 |
| G | 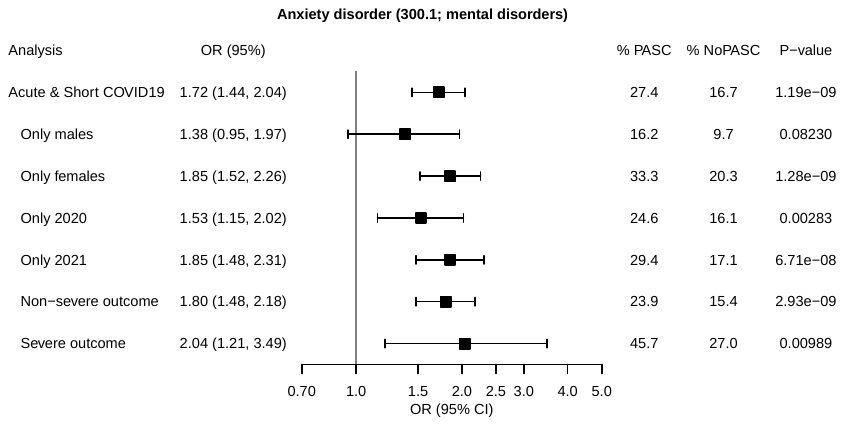 |
| H | 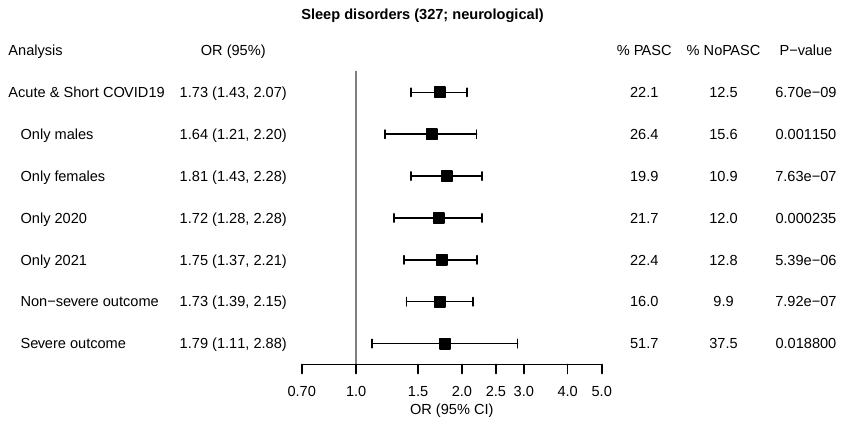 |
| I | 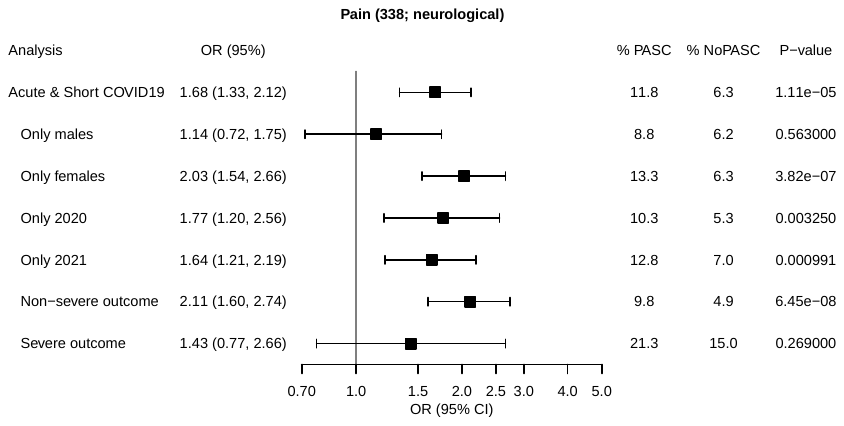 |
| J | 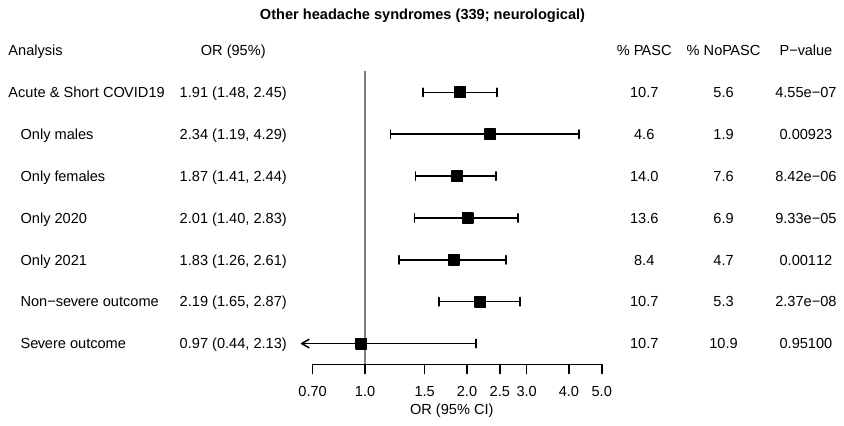 |
| K | 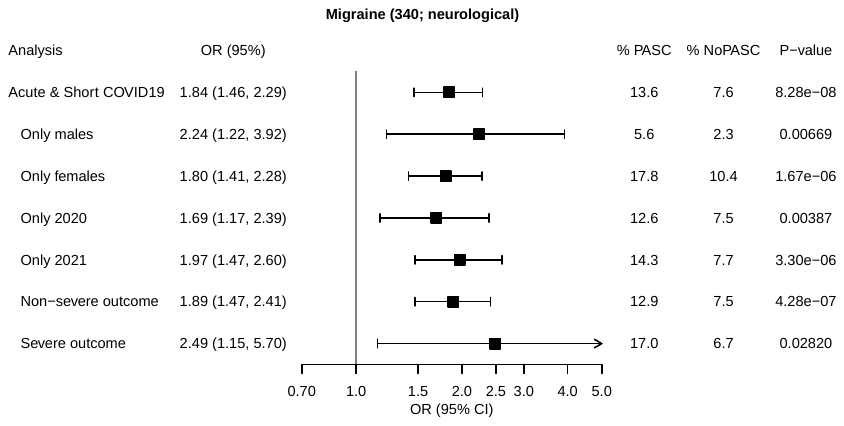 |
| L | 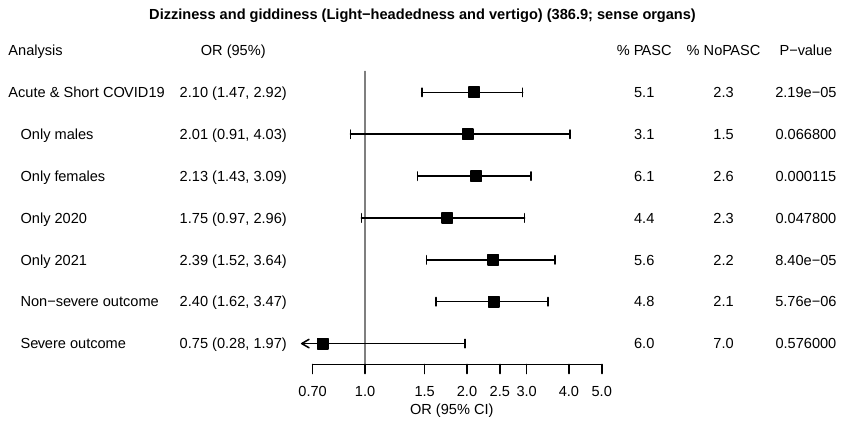 |
| M | 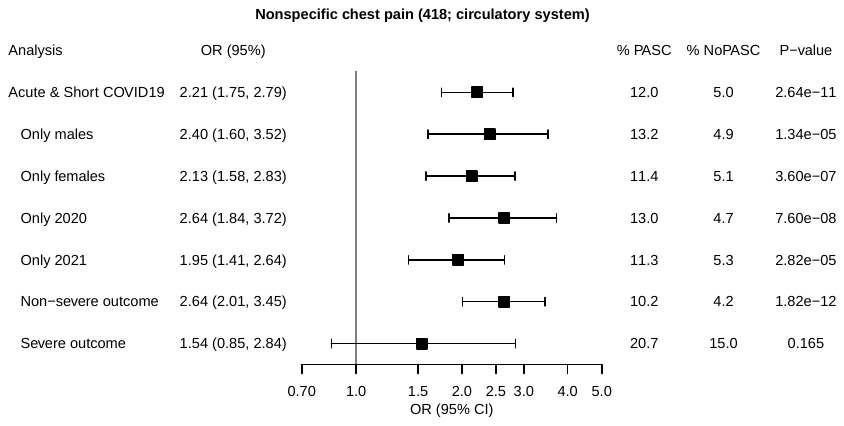 |
| N | 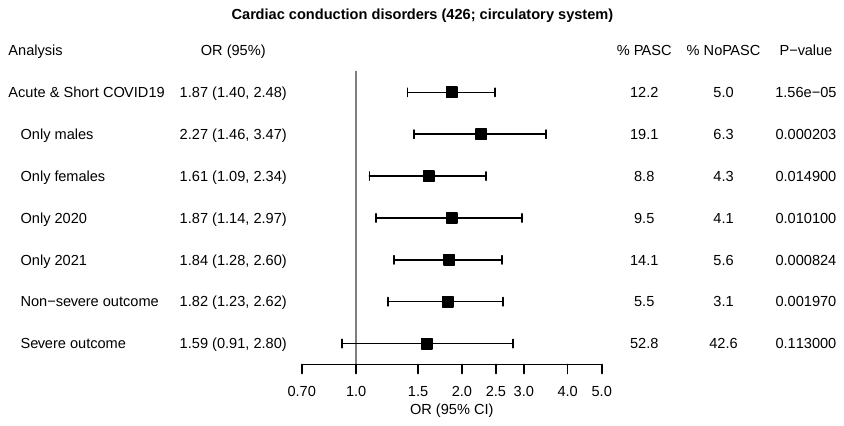 |
| O | 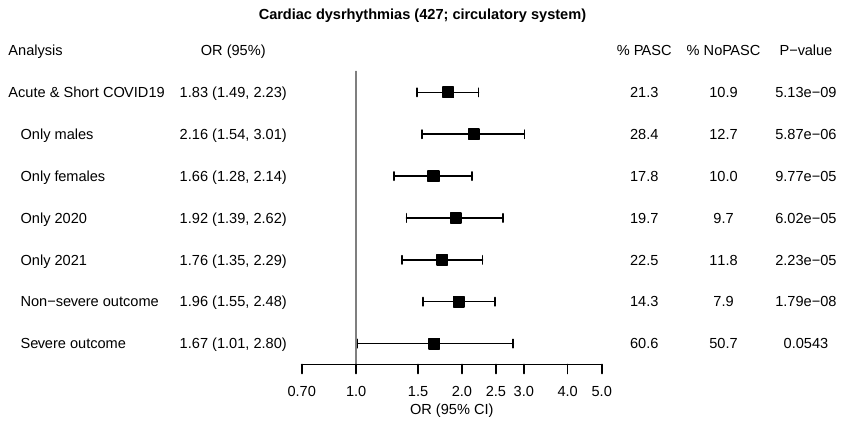 |
| P | 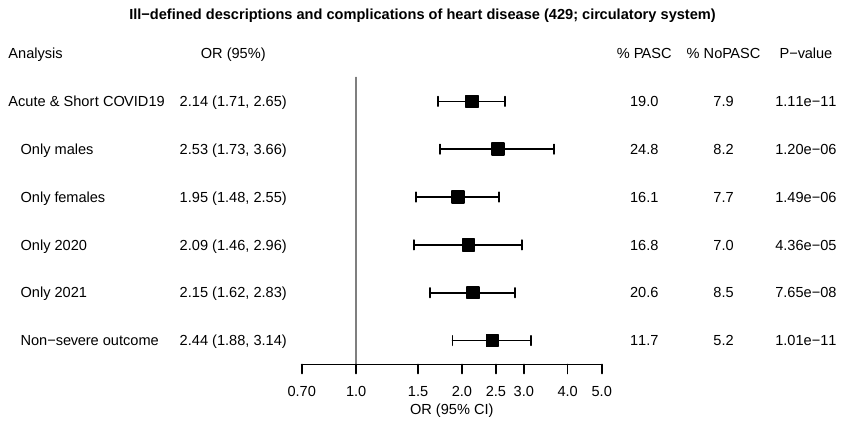 |
| Q | 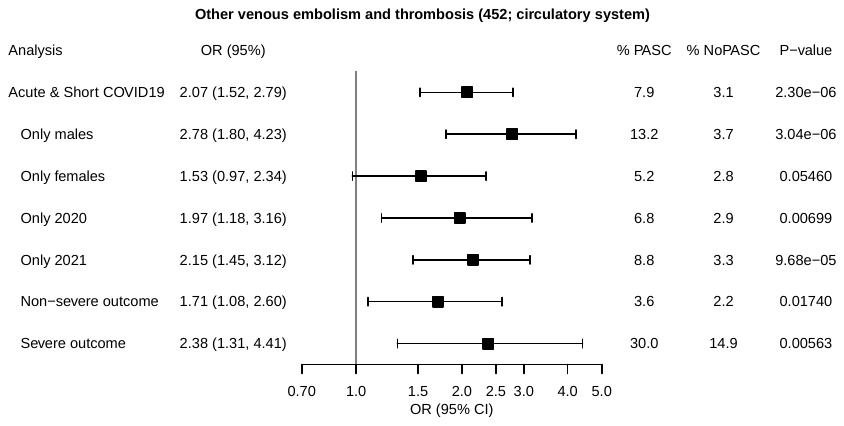 |
| R | 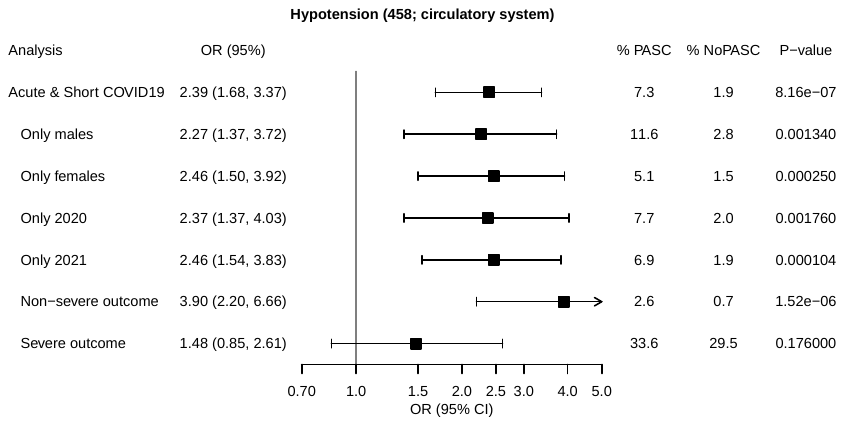 |
| S | 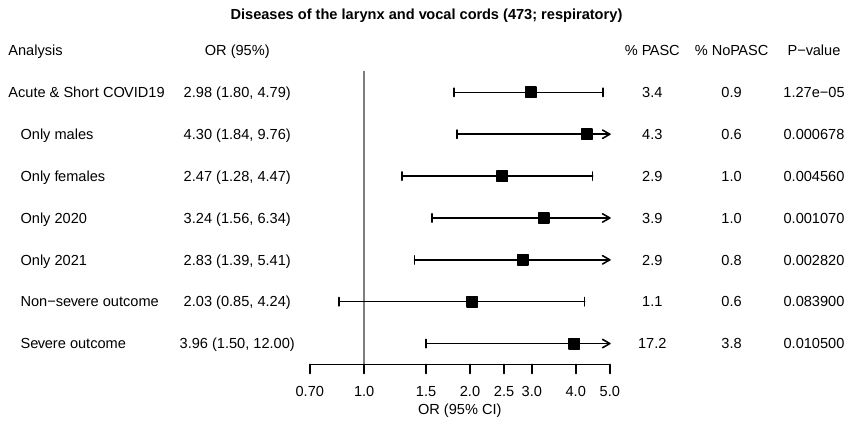 |
| T | 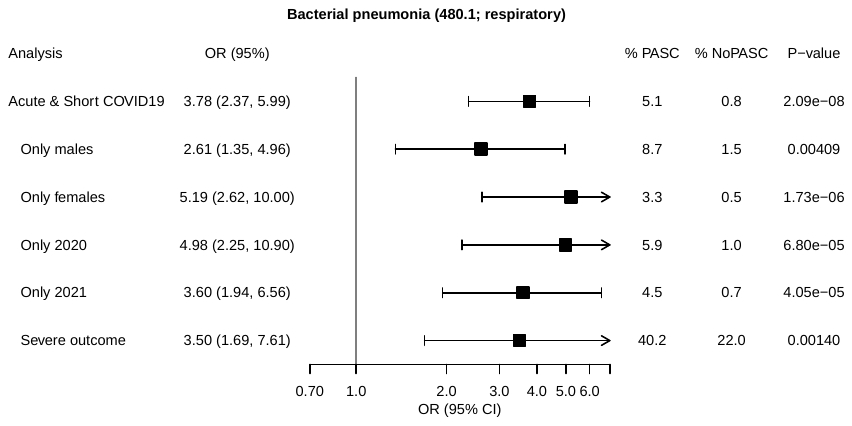 |
| U | 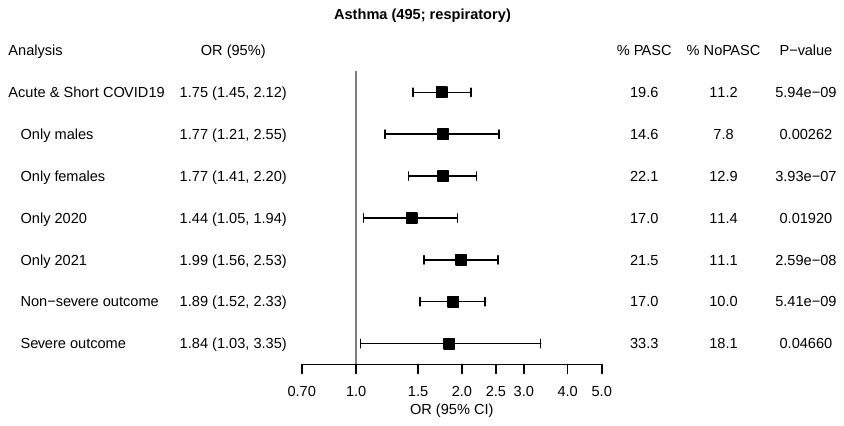 |
| V | 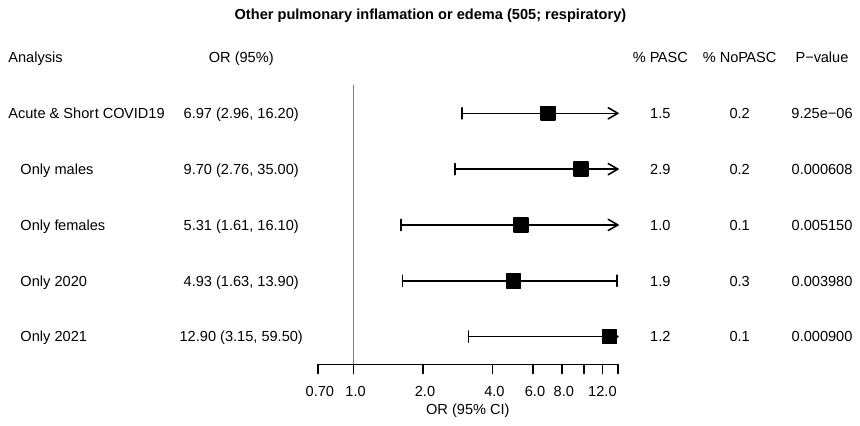 |
| W | 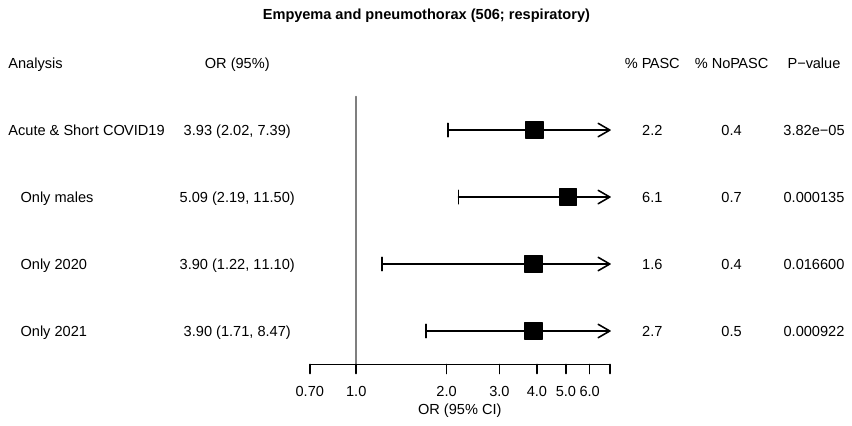 |
| X | 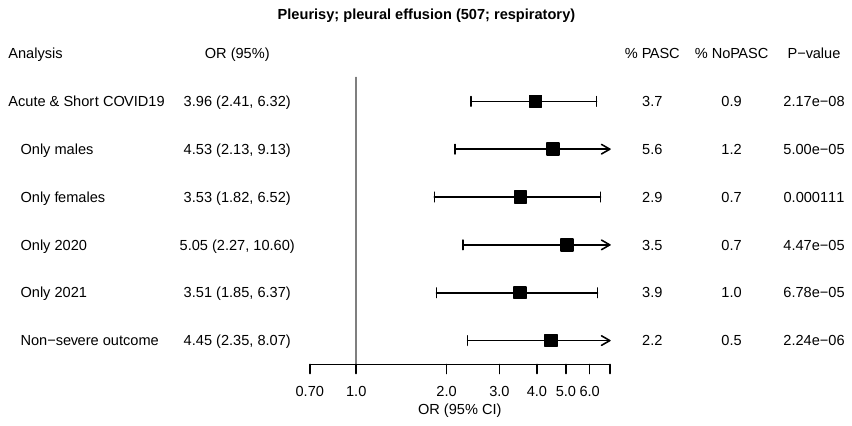 |
| Y | 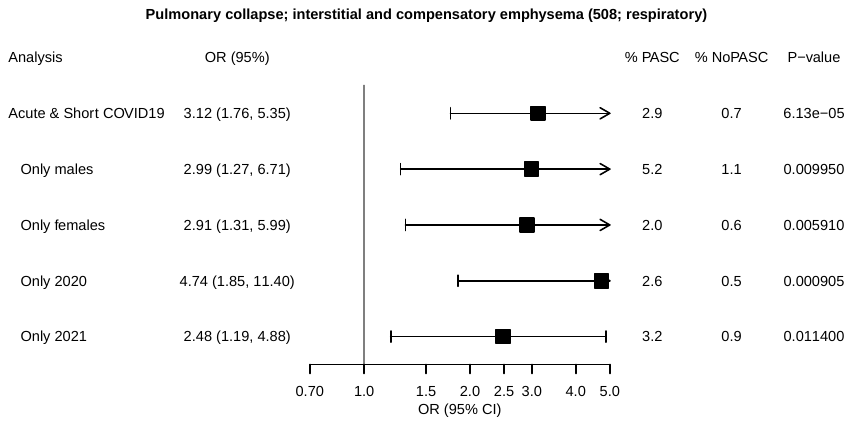 |
| Z | 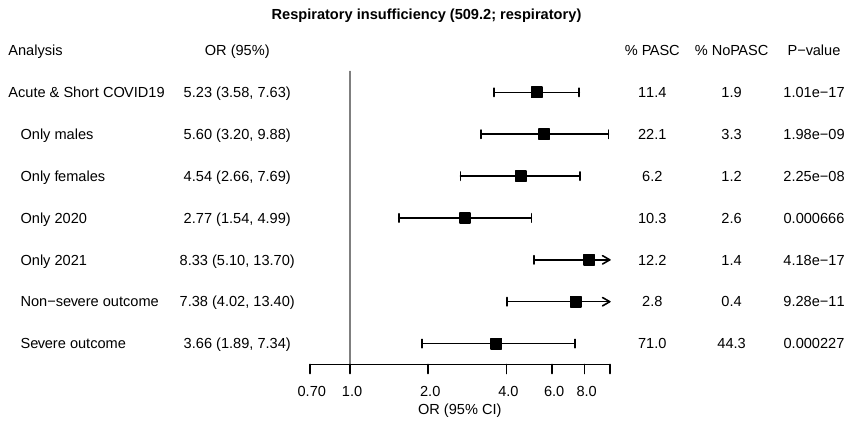 |
| AA | 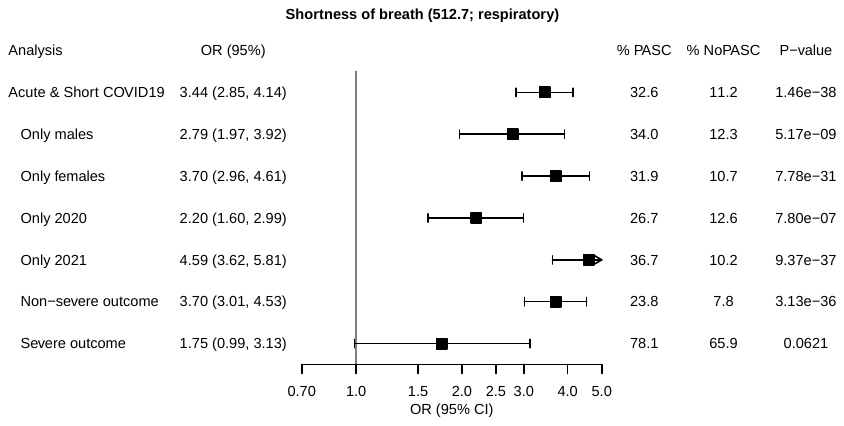 |
| AB | 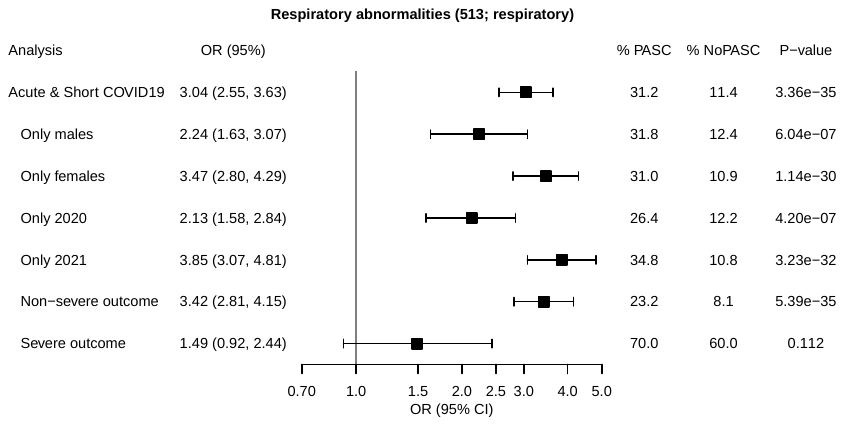 |
| AC | 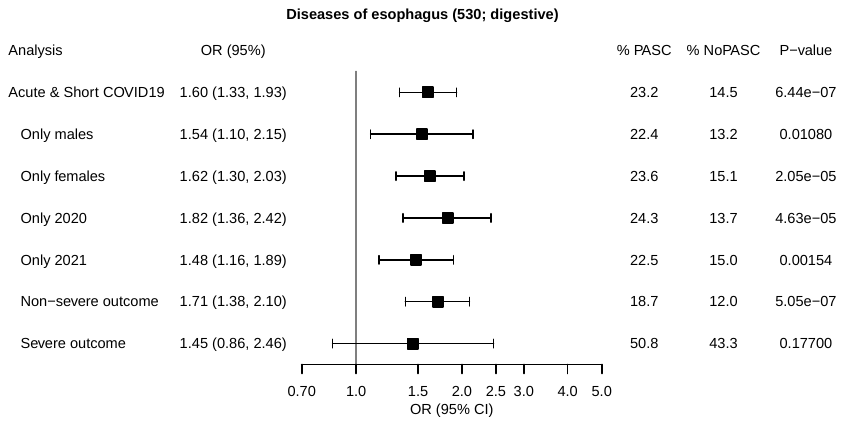 |
| AD | 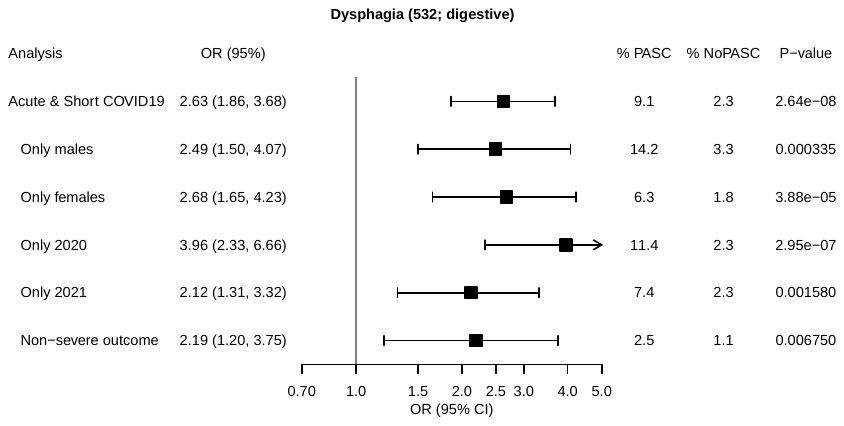 |
| AE | 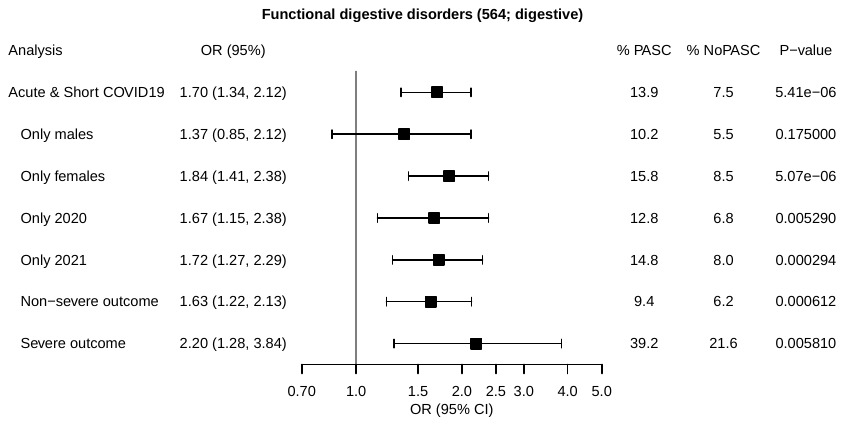 |
| AF | 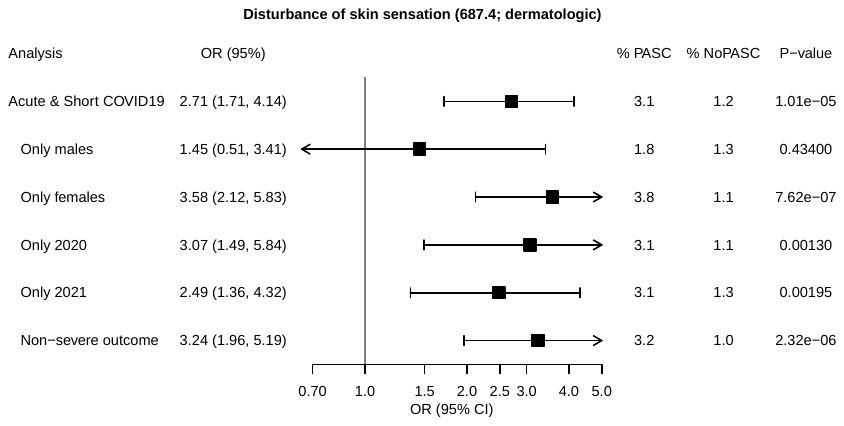 |
| AG | 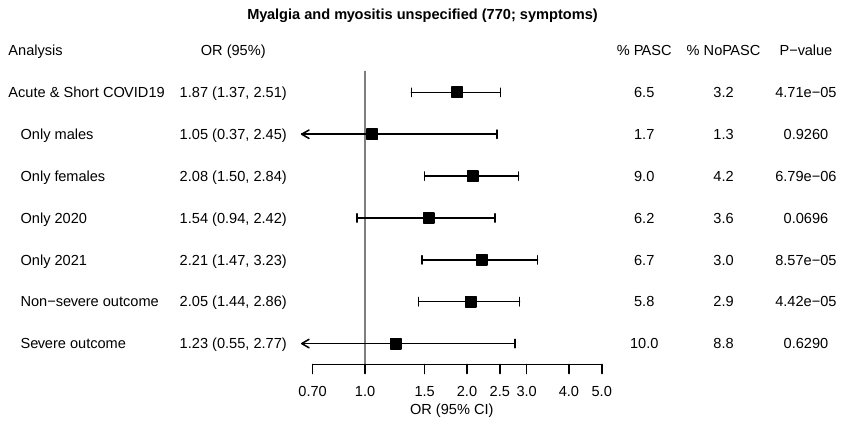 |
| AH | 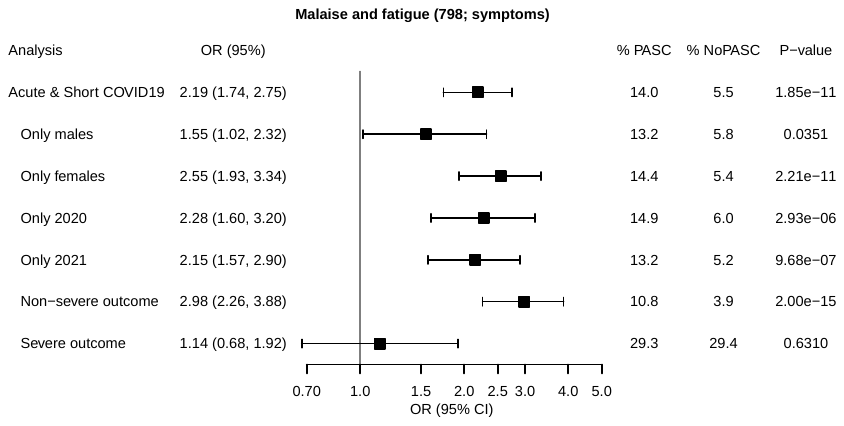 |
| AI | 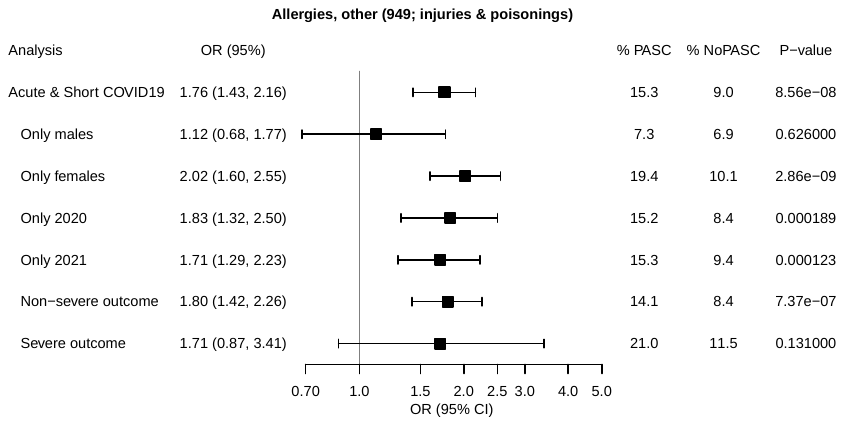 |
| AJ | 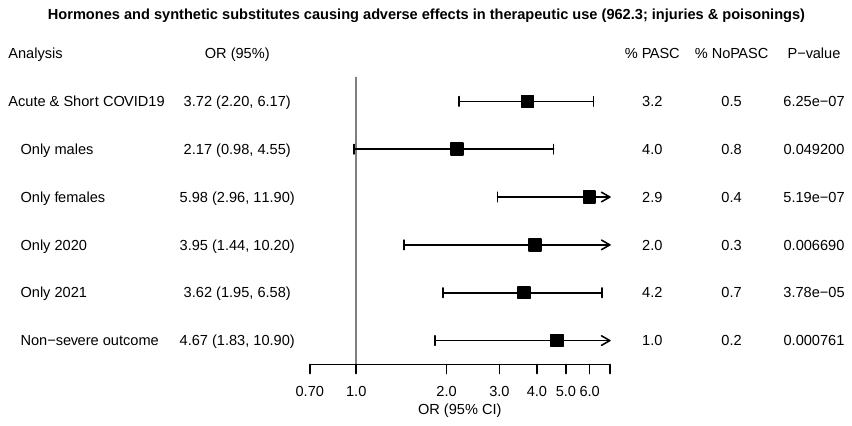 |
| AK | 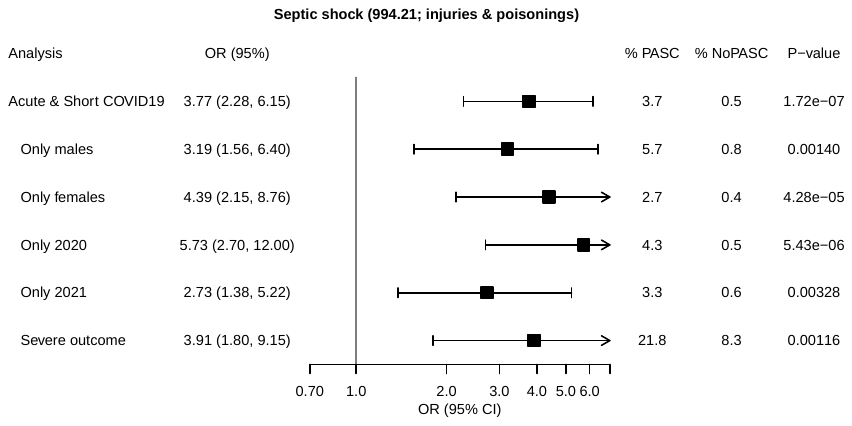 |

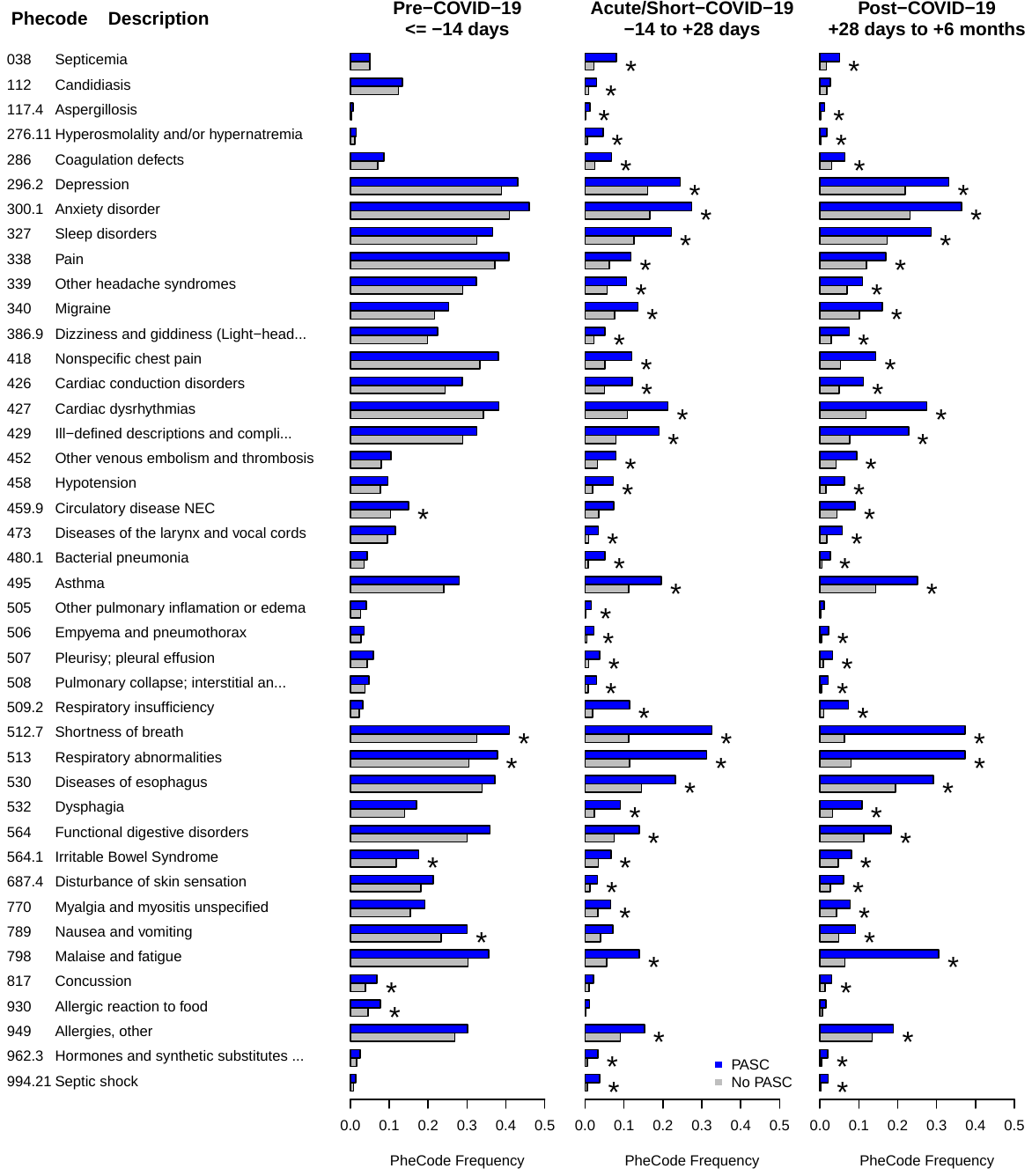

**Figure S5. Comparison of PheCode prevalence during pre-COVID-19, acute/short-COVID-19, and post-COVID-19 periods in cases and controls.** Only PheCodes that were significantly enriched among PASC patients in the pre-COVID-19 and/or acute/short-COVID-19 periods are shown. Of parent/child PheCodes only the phecode with the stronger association signal is shown. PheCodes that were phenome-wide significant in the corresponding PheWAS are indicated with an asterisk. Phenome-wide significant enriched PheCodes are indicated by an asterisk.

**Table S1**: PASC Problem list

| **Problem list description** | **Mapped to ICD10 Code** |
| --- | --- |
| Chronic post-COVID-19 syndrome | B94.8 |
| Shortness of breath with exposure to COVID-19 virus | R06.02 |
| COVID-19 long hauler | U09.9 |
| COVID-19 long hauler manifesting chronic anxiety | U09.9 |
| COVID-19 long hauler manifesting chronic concentration deficit | U09.9 |
| COVID-19 long hauler manifesting chronic cough | U09.9 |
| COVID-19 long hauler manifesting chronic dyspnea | U09.9 |
| COVID-19 long hauler manifesting chronic fatigue | U09.9 |
| COVID-19 long hauler manifesting chronic headache | U09.9 |
| COVID-19 long hauler manifesting chronic joint pain | U09.9 |
| COVID-19 long hauler manifesting chronic loss of smell | U09.9 |
| COVID-19 long hauler manifesting chronic loss of smell and taste | U09.9 |
| COVID-19 long hauler manifesting chronic loss of taste | U09.9 |
| COVID-19 long hauler manifesting chronic muscle pain | U09.9 |
| COVID-19 long hauler manifesting chronic neurologic symptoms | U09.9 |
| COVID-19 long hauler manifesting chronic palpitations | U09.9 |
| Long COVID | U09.9 |
| Multiple persistent symptoms after COVID-19 | U09.9 |
| Persistent dyspnea after COVID-19 | U09.9 |
| Persistent fatigue after COVID-19 | U09.9 |
| Persistent neurologic symptoms after COVID-19 | U09.9 |
| Persistent shortness of breath after COVID-19 | U09.9 |
| Post covid-19 condition, unspecified | U09.9 |
| Post-acute COVID-19 syndrome | U09.9 |
| Post-acute sequelae of COVID-19 (PASC) | U09.9 |
| Post-COVID chronic anxiety | U09.9 |
| Post-COVID chronic concentration deficit | U09.9 |
| Post-COVID chronic cough | U09.9 |
| Post-COVID chronic dyspnea | U09.9 |
| Post-COVID chronic fatigue | U09.9 |
| Post-COVID chronic headache | U09.9 |
| Post-COVID chronic joint pain | U09.9 |
| Post-COVID chronic loss of smell | U09.9 |
| Post-COVID chronic loss of smell and taste | U09.9 |
| Post-COVID chronic loss of taste | U09.9 |
| Post-COVID chronic muscle pain | U09.9 |
| Post-COVID chronic neurologic symptoms | U09.9 |
| Post-COVID chronic palpitations | U09.9 |
| Post-COVID chronic shortness of breath | U09.9 |
| Post-COVID syndrome | U09.9 |
| Post-COVID-19 condition | U09.9 |
| Post-COVID-19 syndrome | U09.9 |
| Post-COVID-19 syndrome manifesting as chronic anxiety | U09.9 |
| Post-COVID-19 syndrome manifesting as chronic concentration deficit | U09.9 |
| Post-COVID-19 syndrome manifesting as chronic cough | U09.9 |
| Post-COVID-19 syndrome manifesting as chronic dyspnea | U09.9 |
| Post-COVID-19 syndrome manifesting as chronic fatigue | U09.9 |
| Post-COVID-19 syndrome manifesting as chronic headache | U09.9 |
| Post-COVID-19 syndrome manifesting as chronic joint pain | U09.9 |
| Post-COVID-19 syndrome manifesting as chronic loss of smell | U09.9 |
| Post-COVID-19 syndrome manifesting as chronic loss of smell and taste | U09.9 |
| Post-COVID-19 syndrome manifesting as chronic loss of taste | U09.9 |
| Post-COVID-19 syndrome manifesting as chronic muscle pain | U09.9 |
| Post-COVID-19 syndrome manifesting as chronic neurologic symptoms | U09.9 |
| Post-COVID-19 syndrome manifesting as chronic palpitations | U09.9 |
| Post-COVID-19 syndrome manifesting as chronic shortness of breath | U09.9 |
| COVID-19 long hauler manifesting chronic decreased mobility and endurance | Z74.09 |
| Post-COVID chronic decreased mobility and endurance | Z74.09 |
| Post-COVID-19 syndrome manifesting as chronic decreased mobility and endurance | Z74.09 |

**Table S2: PASC symptom and concurrent symptom mapping**

| **PheCode** | **PheCode Description** | **PSL**  **Key Words** | **ICD10 Diagnosis mapped to PSL** | **Symptom described in  Chen et al 2022** |
| --- | --- | --- | --- | --- |
| 260.6 | Anorexia | Appetite | R63.0 | Appetite / Eating disorder |
| 292 | Neurological disorders | Neurologic or cognitive deficit | R41 | Memory problems / Concentration / Confusion / Brain fog |
| 296.2 | Depression | Depression | F32 | Depression |
| 300.1 | Anxiety disorder | Anxiety | F41 | Anxiety |
| 327 | Sleep disorders | Sleep apnea | G47 | Sleep problems |
| 338.2 | Chronic pain | Chronic pain | G89.2 | n/a |
| 339 | Any headache syndromes | Headache | R51 | Headache |
| 340 | Migraine | Migraine | G43 | n/a |
| 350.6 | Disturbances of sensation of smell and taste | Smell and taste | R43.8 | Smell or Taste |
| 386.9 | Dizziness and giddiness (Light-headedness and vertigo) | Dizziness | R42 | Dizziness |
| 418 | Nonspecific chest pain | Chest pain | R07 | Chest pain |
| 427.7 | Tachycardia NOS | Tachycardia | R00 | Tachycardia |
| 427.9 | Palpitations | Palpitations | R00 | n/a |
| 465.2 | Acute pharyngitis | Sore throat | J02.9 | Sore throat |
| 473.4 | Voice disturbance | Dysphonia | R49.0 | n/a |
| 495 | Asthma | Asthma | J45 | n/a |
| 496 | Chronic airway obstruction | COPD | J44 | n/a |
| 512.7 | Shortness of breath | Dyspnea | R06.0 | Dyspnea |
| 512.8 | Cough | Cough | R05 | Cough |
| 561 | Symptoms involving digestive system | Diarrhea | R19.7 | Diarrhea |
| 687.1 | Rash and other nonspecific skin eruption | Rash | R21 | n/a |
| 704.1 | Alopecia | Hair loss | L65 | Hair loss |
| 745 | Pain in joint | Joint pain | M25.5 | Joint pain |
| 760 | Back pain | Back pain | M54 | n/a |
| 770 | Myalgia and myositis unspecified | Myalgia | M79.1 | Myalgia |
| 772.3 | Muscle weakness | Muscle weakness | M62.81 | n/a |
| 783 | Fever of unknown origin | Fever | R50 | Fever |
| 785 | Abdominal pain | Abdominal pain | R10 | Abdominal pain |
| 798 | Malaise and fatigue | Fatigue | R53 | Fatigue |

**Table S3**. Covariate summary and missingness in the unmatched and matched cohort (see **Figure 2**)

| ***Standard Set of Covariates*** |  | | **Missingness, n (%)** | |
| --- | --- | --- | --- | --- |
| **Covariate** | **Data Type** | | **Unmatched cohort n = 63,675** | **Matched cohort n = 18,929** |
| Age at positive COVID-19 test/diagnosis | Continuous | | 0 (0) | 0 (0)^e^ |
| Gender | Binary | | 5 (0) | 0 (0)^e^ |
| Self-reported Race / Ethnicity ^a^ | Categorical | | 0 (0) | 0 (0)^e^ |
| Elixhauser Score (AHRQ) | Continuous | | 0 (0) | 0 (0) |
| NDI | Categorical (quartiles) | | 5,793 (9.1) | 1,335 (7.1) |
| Population density | Categorical (quartiles) | | 5,793 (9.1) | 1,335 (7.1) |
| Health Care Worker Status | Binary | | 0 (0)^f^ | 0 (0)^f^ |
| Severity ^b^ | Binary | | 0 (0)^f^ | 0 (0)^f^ |
| Vaccination status at time of test ^c^ | Categorical | | 0 (0)^f^ | 0 (0)^f^ |
| ***Analysis dependent Covariates*** | | |  |  |
| **Analysis** | **Covariate** | **Data Type** | **Missingness, n (%)** | |
| Pre-Pandemic PheWAS | Pre-Pandemic Years in EHR | Continuous | 1,360 (2.1) | 99 (0.5) |
| Pre-COVID-19 PheWAS | Pre-Test Years in EHR | Continuous | 0 (0) | 0 (0)^e^ |
| Acute & Short COVID-19 PheWAS | Post-COVID-19-Test Years in EHR | Continuous | 0 (0) | 0 (0)^e^ |
| Post-COVID-19 Symptom PheWAS | Post-COVID-19-Test Years in EHR | Continuous | 0 (0) | 0 (0)^e^ |

^a^ Self-reported Race / Ethnicity factor levels: Caucasian/Non-Hispanic, African American/Non-Hispanic, Other Race or Ethnicity, and Unknown Race or Ethnicity

^b^ mild: asymptomatic/non-hospitalized; severe: hospitalized, ICU stay or deceased

^c^ unvaccinated, partially vaccinated, fully vaccinated, boosted

^e^ complete data was requirement for matching

^f^ based on available EHR documentation (“documented” versus “not documented or unknown”); actual missingness unknown

**Table S4.** Main and sensitivity PheWAS. Each analysis only includes individual who were COVID-19 positive for the first time before 2022.

| **Analysis** | **Phenome Description (relative to index date)** | **n  PASC** | **n  No PASC (Matched)** | **n PheCodes*** |
| --- | --- | --- | --- | --- |
| **Main Analysis** | Pre-existing conditions / pre-COVID-19 symptoms (before -14 days) | 1,212 | 11,919 | 1,405 |
| **Sensitivity Analyses** | Females only | 793 | 7,882 | 1,237 |
|  | Males only | 419 | 4,037 | 928 |
|  | 1. Positive in 2020 | 381 | 3,671 | 1,013 |
|  | 1. Positive in 2021 | 831 | 8,248 | 1,285 |
|  | Asymptomatic / mild COVID-19 outcome | 1,009 | 9,393 | 1,273 |
|  | Severe COVID-19 outcome | 203 | 204 | 944 |
|  | Max 2 years before COVID-19 test | 1,212 | 11,919 | 1,061 |
|  | Pre-existing conditions before pandemic (before 2020) | 1,085 | 10,533 | 1,347 |
| **Main Analysis** | Acute short COVID-19 period symptoms (between -14 and + 28 days) | 874 | 8,671 | 664 |
| **Sensitivity Analyses** | Females only | 578 | 5,766 | 510 |
|  | Males only | 296 | 2,905 | 365 |
|  | 1. Positive in 2020 | 368 | 3,623 | 397 |
|  | 2. Positive in 2021 | 506 | 5,048 | 504 |
|  | Asymptomatic / Mild COVID-19 | 724 | 6,799 | 454 |
|  | Severe COVID-19 | 150 | 160 | 419 |
| **Main Analysis** | Post-COVID-19 symptoms (between +28 days and +6 months) | 1,256 | 12,492 | 961 |

* PheCode prevalence >= 5 among individuals with and without PASC (complete case analysis)

**Table S5**: Concurrent diagnoses on day of the first PASC diagnosis. Only diagnoses that matched the 24 listed categories are shown.

| **Symptoms** | **n** | **% with Unspecified PASC (n = 1,724)** | **% Without Unspecified PASC (n = 1,362)** |
| --- | --- | --- | --- |
| Shortness of breath | 469 | 27.2 | 34.4 |
| Anxiety disorder | 417 | 24.2 | 30.6 |
| Malaise and fatigue | 388 | 22.5 | 28.5 |
| Depression | 370 | 21.5 | 27.2 |
| Sleep disorders | 346 | 20.1 | 25.4 |
| Asthma | 321 | 18.6 | 23.6 |
| Other headache syndromes | 291 | 16.9 | 21.4 |
| Migraine | 188 | 10.9 | 13.8 |
| Cough | 177 | 10.3 | 13.0 |
| Pain in joint | 172 | 10.0 | 12.6 |
| Back pain | 158 | 9.2 | 11.6 |
| Neurological disorders | 155 | 9.0 | 11.4 |
| Nonspecific chest pain | 143 | 8.3 | 10.5 |
| Chronic pain | 132 | 7.7 | 9.7 |
| Tachycardia NOS | 114 | 6.6 | 8.4 |
| Myalgia and myositis unspecified | 115 | 6.7 | 8.4 |
| Chronic airway obstruction | 101 | 5.9 | 7.4 |
| Symptoms involving digestive system | 100 | 5.8 | 7.3 |
| Abdominal pain | 91 | 5.3 | 6.7 |
| Palpitations | 66 | 3.8 | 4.8 |
| Rash and other nonspecific skin eruption | 65 | 3.8 | 4.8 |
| Dizziness and giddiness (Light-headedness and vertigo) | 64 | 3.7 | 4.7 |
| Fever of unknown origin | 46 | 2.7 | 3.4 |
| Disturbances of sensation of smell and taste | 44 | 2.6 | 3.2 |
| Voice disturbance | 23 | 1.3 | 1.7 |
| Muscle weakness | 15 | 0.9 | 1.1 |
| Acute pharyngitis | 12 | 0.7 | 0.9 |
| Anorexia | 11 | 0.6 | 0.8 |
| Alopecia | 8 | 0.5 | 0.6 |
| Unspecified PASC | 362 | 21.0 | 0.0 |

**Table S6.** Enrichment 29 known PASC symptoms among post-COVID-19 diagnoses (observed between +28 days and 6 months after being COVID-19 positive) in PASC cases compared to “No PASC” controls

| **PheCode** | **PheCode Description** | **PheCode Category** | **PheCode Frequeny [PASC]** | **PheCode Frequency [No PASC]** | **OR (95% CI)** | **P-value** | **Significance Level** |
| --- | --- | --- | --- | --- | --- | --- | --- |
| 512.7 | Shortness of breath | respiratory | 425 / 1139 (37.3) | 744 / 11752 ( 6.3) | 9.03 (7.77, 10.5) | 2.94E-181 | phenome-wide |
| 798 | Malaise and fatigue | symptoms | 384 / 1256 (30.6) | 803 / 12492 ( 6.4) | 6.17 (5.33, 7.14) | 2.32E-132 | phenome-wide |
| 512.8 | Cough | respiratory | 136 / 850 (16.0) | 457 / 11465 ( 4.0) | 4.39 (3.54, 5.41) | 1.61E-42 | phenome-wide |
| 292 | Neurological disorders | mental disorders | 160 / 1228 (13.0) | 439 / 12274 ( 3.6) | 3.57 (2.91, 4.35) | 8.85E-36 | phenome-wide |
| 427.7 | Tachycardia NOS | circulatory system | 118 / 1003 (11.8) | 286 / 11112 ( 2.6) | 4.39 (3.45, 5.56) | 5.58E-34 | phenome-wide |
| 418 | Nonspecific chest pain | circulatory system | 180 / 1253 (14.4) | 660 / 12487 ( 5.3) | 2.87 (2.39, 3.44) | 2.98E-30 | phenome-wide |
| 427.9 | Palpitations | circulatory system | 97 / 982 ( 9.9) | 306 / 11132 ( 2.7) | 3.88 (3.03, 4.92) | 2.02E-28 | phenome-wide |
| 300.1 | Anxiety disorder | mental disorders | 406 / 1114 (36.4) | 2628 / 11368 (23.1) | 1.95 (1.70, 2.23) | 4.55E-22 | phenome-wide |
| 350.6 | Disturbances of sensation of smell and taste | neurological | < 50 / 1172 ( 3.0) | < 50 / 12091 ( 0.4) | 7.69 (4.93, 11.9) | 4.51E-21 | phenome-wide |
| 327 | Sleep disorders | neurological | 358 / 1253 (28.6) | 2152 / 12478 (17.2) | 1.92 (1.67, 2.20) | 1.11E-20 | phenome-wide |
| 495 | Asthma | respiratory | 300 / 1196 (25.1) | 1742 / 12171 (14.3) | 1.98 (1.72, 2.29) | 1.36E-20 | phenome-wide |
| 386.9 | Dizziness and giddiness (Light-headedness and vertigo) | sense organs | 94 / 1252 ( 7.5) | 367 / 12434 ( 3.0) | 2.68 (2.10, 3.39) | 3.65E-16 | phenome-wide |
| 296.2 | Depression | mental disorders | 351 / 1059 (33.1) | 2448 / 11188 (21.9) | 1.79 (1.55, 2.07) | 8.83E-16 | phenome-wide |
| 783 | Fever of unknown origin | symptoms | 73 / 1256 ( 5.8) | 196 / 12492 ( 1.6) | 3.15 (2.34, 4.19) | 6.41E-15 | phenome-wide |
| 473.4 | Voice disturbance | respiratory | < 50 / 958 ( 4.3) | 113 / 10361 ( 1.1) | 3.43 (2.33, 4.94) | 5.86E-11 | phenome-wide |
| 340 | Migraine | neurological | 190 / 1187 (16.0) | 1224 / 12007 (10.2) | 1.75 (1.47, 2.08) | 1.74E-10 | phenome-wide |
| 770 | Myalgia and myositis unspecified | symptoms | 97 / 1256 ( 7.7) | 532 / 12492 ( 4.3) | 1.87 (1.48, 2.35) | 9.22E-08 | phenome-wide |
| 785 | Abdominal pain | symptoms | 159 / 1256 (12.7) | 1003 / 12492 ( 8.0) | 1.61 (1.34, 1.93) | 2.80E-07 | phenome-wide |
| 760 | Back pain | symptoms | 225 / 1256 (17.9) | 1555 / 12492 (12.4) | 1.51 (1.28, 1.76) | 4.45E-07 | phenome-wide |
| 496 | Chronic airway obstruction | respiratory | 92 / 988 ( 9.3) | 507 / 10936 ( 4.6) | 1.93 (1.49, 2.49) | 4.92E-07 | phenome-wide |
| 745 | Pain in joint | musculoskeletal | 219 / 1256 (17.4) | 1537 / 12492 (12.3) | 1.51 (1.28, 1.77) | 5.62E-07 | phenome-wide |
| 772.3 | Muscle weakness | symptoms | < 50 / 1221 ( 2.9) | 103 / 12252 ( 0.8) | 2.67 (1.76, 3.95) | 1.11E-06 | phenome-wide |
| 704.1 | Alopecia | dermatologic | < 50 / 1238 ( 3.5) | 177 / 12339 ( 1.4) | 2.31 (1.62, 3.22) | 1.39E-06 | phenome-wide |
| 339 | Other headache syndromes | neurological | 122 / 1119 (10.9) | 808 / 11591 ( 7.0) | 1.65 (1.34, 2.03) | 2.21E-06 | phenome-wide |
| 465.2 | Acute pharyngitis | respiratory | < 50 / 1221 ( 2.8) | 162 / 12232 ( 1.3) | 2.27 (1.53, 3.28) | 1.66E-05 | phenome-wide |
| 338.2 | Chronic pain | neurological | 164 / 1207 (13.6) | 1122 / 12121 ( 9.3) | 1.49 (1.24, 1.78) | 1.87E-05 | phenome-wide |
| 687.1 | Rash and other nonspecific skin eruption | dermatologic | 78 / 1183 ( 6.6) | 472 / 12148 ( 3.9) | 1.71 (1.32, 2.19) | 2.71E-05 | phenome-wide |
| 561 | Symptoms involving digestive system | digestive | < 50 / 981 ( 1.9) | 139 / 10676 ( 1.3) | 1.56 (0.93, 2.47) | 0.069 | n.s. |
| 260.6 | Anorexia | endocrine/metabolic | < 50 / 1047 ( 0.8) | < 50 / 11384 ( 0.4) | 1.81 (0.79, 3.70) | 0.106 | n.s. |

Notes: phenome-wide: P < 0.05/961 tested PheCodes (P < 5.1E-05); n.s.: not significant

**Table S7.** PheRS Evaluation in the testing data (COVID-19 positive in 2022). PheRS1* was based on phecodes that reached P < 1-E3 in the PheWAS with the pre-COVID-19 training data (1,256 cases and 11,674 controls; COVID-19 positive in 2020/2021) while PheRS2* was based on phecodes that reached P < 1-E3 in the PheWAS with the acute & short COVID-19 training data (874 cases and 8,144 controls; COVID-19 positive in 2020/2021 & at least 28 days between first COVID-19 and first PASC diagnosis). Underlying weights can be found in **File S1J and Table S8**.

| **Predictor** | **Testing Data** | | **AAUC ^a^**  **95% CI** | **Pseudo-R^2 b^** | **Brier Score** |
| --- | --- | --- | --- | --- | --- |
|  | **n Cases** | **n Controls** |  |  |  |
| PheRS1* | 349 | 3248 | 0.545 (0.514, 0.576) | n/a ^c^ | n/a ^c^ |
| PheRS1* | 123 | 1154 | 0.551 (0.495, 0.611) | 0.0086 | 0.0857 |
| PheRS2* |  |  | 0.595 (0.540, 0.653) | 0.0439 | 0.0830 |
| PheRS1 & PheRS2* |  |  | 0.601 (0.548, 0.658) | 0.0435 | 0.0831 |

^a^ Adjusted for age at index date, gender, race/ethnicity, Elixhauser Score, population density, NDI, health care worker status, vaccination status, pre-test years in EHR, and severity

^b^ Nagelkerke [Cragg and Uhler])

^c^ not applicable, only useful in evaluating multiple models predicting the same outcome on the same dataset

**Table S8** Weights for combining PheRS1 and PheRS2 or PheRS1* and PheRS2*. PheRS were standardized (mean =0 and sd = 1) before calculating the weights and before calculating the combined PheRSs.

| **PheRS Combination** | **Predictor** | **Weight*** |
| --- | --- | --- |
| PheRS1 & PheRS2 Combined | PheRS1 | 0.08289 |
|  | PheRS2 | 0.52371 |
| PheRS1* & PheRS2* Combined | PheRS1* | 0.08267 |
|  | PheRS2* | 0.51564 |
